# Deep learning-derived splenic radiomics, genomics, and coronary artery disease

**DOI:** 10.1101/2024.08.16.24312129

**Authors:** Meghana Kamineni, Vineet Raghu, Buu Truong, Ahmed Alaa, Art Schuermans, Sam Friedman, Christopher Reeder, Romit Bhattacharya, Peter Libby, Patrick T. Ellinor, Mahnaz Maddah, Anthony Philippakis, Whitney Hornsby, Zhi Yu, Pradeep Natarajan

## Abstract

**Background:** Despite advances in managing traditional risk factors, coronary artery disease (CAD) remains the leading cause of mortality. Circulating hematopoietic cells influence risk for CAD, but the role of a key regulating organ, spleen, is unknown. The understudied spleen is a 3-dimensional structure of the hematopoietic system optimally suited for unbiased radiologic investigations toward novel mechanistic insights.

**Methods:** Deep learning-based image segmentation and radiomics techniques were utilized to extract splenic radiomic features from abdominal MRIs of 42,059 UK Biobank participants. Regression analysis was used to identify splenic radiomics features associated with CAD. Genome-wide association analyses were applied to identify loci associated with these radiomics features. Overlap between loci associated with CAD and the splenic radiomics features was explored to understand the underlying genetic mechanisms of the role of the spleen in CAD.

**Results:** We extracted 107 splenic radiomics features from abdominal MRIs, and of these, 10 features were associated with CAD. Genome-wide association analysis of CAD-associated features identified 219 loci, including 35 previously reported CAD loci, 7 of which were not associated with conventional CAD risk factors. Notably, variants at 9p21 were associated with splenic features such as run length non-uniformity.

**Conclusions:** Our study, combining deep learning with genomics, presents a new framework to uncover the splenic axis of CAD. Notably, our study provides evidence for the underlying genetic connection between the spleen as a candidate causal tissue-type and CAD with insight into the mechanisms of 9p21, whose mechanism is still elusive despite its initial discovery in 2007. More broadly, our study provides a unique application of deep learning radiomics to non-invasively find associations between imaging, genetics, and clinical outcomes.

## Introduction

Despite advances in the management of traditional risk factors, coronary artery disease (CAD) remains the leading cause of mortality and disability-adjusted life-years (DALYs) worldwide.^1,2^ Advances in CAD prevention beyond targeting traditional risk factors continue to remain limited due to poor understanding and limited mechanistic frameworks of such distinct CAD pathways.

The hematopoietic system has long been known to contribute to CAD, largely through inflammatory cells in both atherogenesis and atherosclerotic cardiovascular disease events.^3^ Inflammation markers, such as high-sensitivity C-reactive protein (hsCRP), are independently predictive of CAD risk. Among individuals with CAD and high hsCRP, a monoclonal antibody targeting interleukin (IL)-1B reduced the risk for recurrent CAD events but increased risk for serious infections.^4^ While this trial validated the causal role of inflammatory cytokines for CAD, the optimal strategy to modulate hematopoietic cells and their products toward CAD risk reduction remains poorly understood.^5^

Longstanding circumstantial evidence has suggested involvement of the spleen, an extramedullary hematopoietic organ, in CAD.^6^ U.S. veterans who underwent splenectomy for trauma during World War II had greater mortality due to CAD in long-term follow-up.^7^ More recently, the spleen was described as an important reservoir for undifferentiated inflammatory myeloid cells that are mobilized in the context of myocardial ischemic injury infiltrating myocardium in murine models.^8^ Myelopoiesis after splenic activation, including during myocardial infarction, further leads to atherosclerosis instability in mice.^9^ Post-mortem human samples from varying times after myocardial infarction demonstrate splenic monocyte depletion early after myocardial infarction, invoking their mobilization early in the event.^10^ (18)F-fluorodeoxyglucose ((18)FDG)-positron emission tomography among patients who sustained acute coronary syndromes showed that increased splenic metabolic activity strongly predicted recurrence.^11^ More recent human genome-wide association studies (GWAS) of CAD have implicated splenic gene regulation. Individual inflammatory genes, including *CCR5*, prioritized through this approach are strongly expressed in the spleen.^12^ Among the top signals for CAD GWAS, splenic tissue is one of the top three tissues enriched for variants residing within strong enhancers and active promoters. However, there is limited understanding regarding the critical factors regulating splenic function in relation to CAD risk.

Advancements in machine learning applied to medical imaging offer new opportunities for unbiased, scalable detection and quantification of subtle alterations in internal organs, including the spleen, where specific circulating biomarkers may be unavailable. Deep learning enables large-scale automatic segmentation of organs in medical images, bypassing time-consuming manual segmentation. Radiomics, an emerging field, quantifies features extracted from these segmentations to offer non-invasive insights into underlying pathologies. These features encapsulate a variety of metrics, such as shape, size, and texture.^13^ For the spleen, radiomics have been used to diagnose and differentiate lymphoma subtypes and predict the recurrence of hepatocellular carcinoma.^14^ Radiomics offers an opportunity to glean novel insights about splenic anatomy as typically only splenic size is annotated in clinical scans.

In this study, we leveraged deep learning and radiomic analyses to extract and discover CAD-relevant splenic features from abdominal magnetic resonance imaging (MRI). Additionally using genomics, we further prioritize previously poorly known CAD-associated loci and genes with key splenic radiomic features. Utilizing a multi-disciplinary approach that integrates advanced imaging analyses, genomics, and clinical outcomes, our study introduces a new framework for understanding the spleen’s potential role in residual CAD risk.

## Methods

### Cohort selection and workflow

The UK Biobank is a volunteer cohort of approximately 500,000 participants aged 40-69 years recruited from 2006 to 2010 with ongoing prospective follow-up.^15^ At baseline, participants provided surveys, biospecimens, anthropometrics, vital signs, and other study-specific procedures. Approximately 50,000 MRIs were performed for a subset of participants after reinvitation beginning in 2014. We limited our study population to those who had abdominal MRIs acquired during the study and whose spleen and liver segments were identifiable after applying our segmentation algorithm. Analysis of the UK Biobank data was approved by the UK Biobank application 7089 and Massachusetts General Hospital IRB protocol 2021P002228. The inclusion and exclusion criteria are visualized in **Supplemental Figure 1**.

Figure 1 illustrates the study workflow. First, we segmented the spleen from abdominal MRIs and extracted comprehensive radiomic features linked to intrinsic splenic properties. Next, we used regression models to discover independent splenic features associated with CAD, which we investigated in subsequent analyses. We then performed GWAS to identify genetic variants associated with each of the CAD-associated splenic phenotypes, building on which we (1) prioritized genes that are likely to be causal and probed their functional relevance to CAD and (2) identified overlapping genetic variants that are significantly associated with both splenic phenotypes and CAD, whose corresponding functions may be the link between the spleen and residual CAD risk.

**Figure 1.**
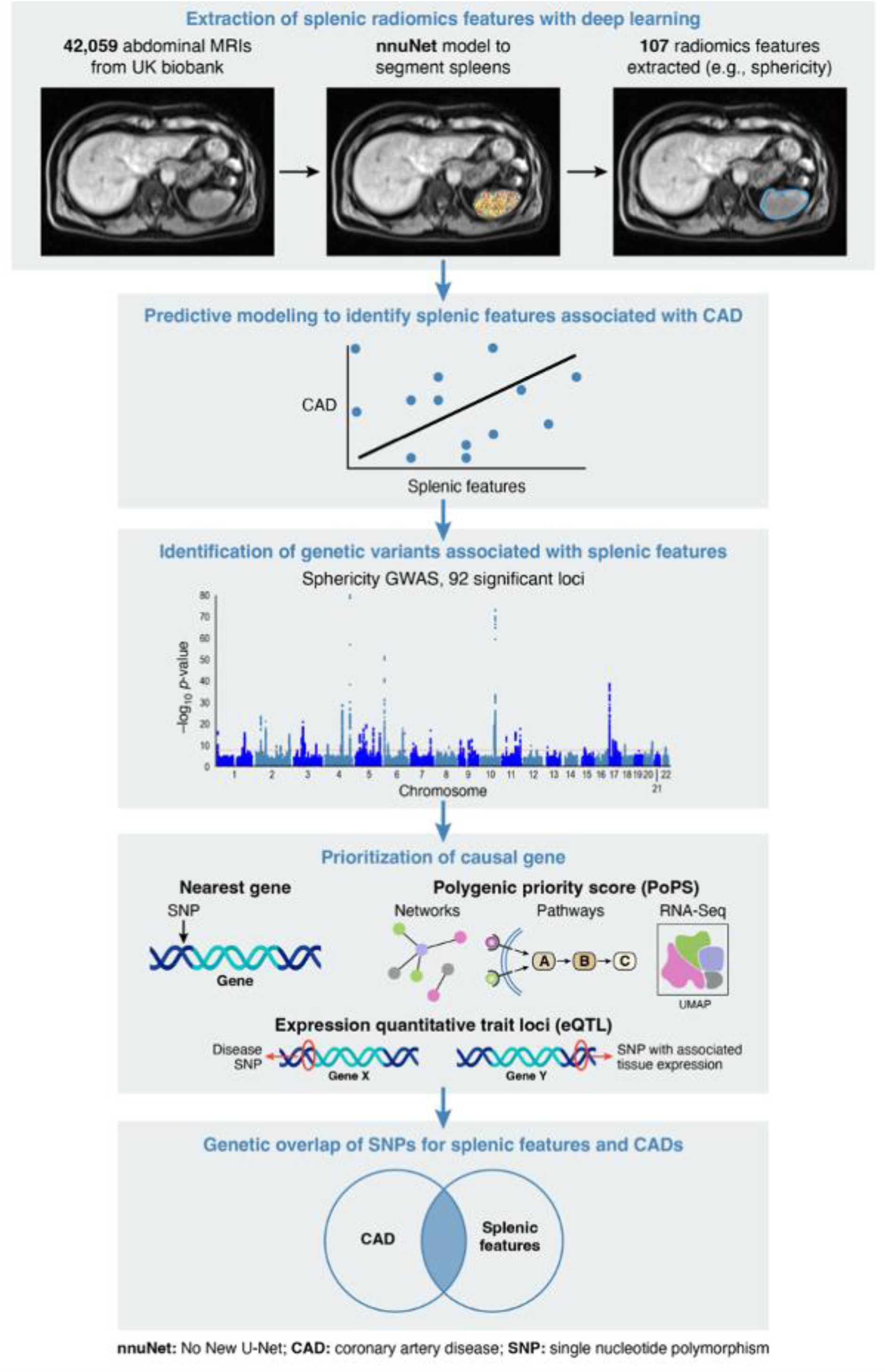
Summary of the workflow to identify splenic features associated with CAD and discover genetic associations. First, radiomics features describing the spleen are extracted from 42,543 abdominal MRIs from the UK Biobank. Second, predictive models of each splenic radiomic feature for CAD are implemented. Genome-wide association studies are then conducted to identify genetic variants significantly associated with CAD-associated splenic radiomic features. Based on the identified genetic variants, genes are then prioritized for further investigation based on three different prioritization techniques: nearest gene, PoPS, and eQTL. Finally, the genetic variants that were associated with splenic radiomics were investigated for association with CAD using summary-level CAD GWAS meta-analysis. PoPs, polygenic priority score. eQTL, expression quantitative trait loci. nnuNet, No New U-Net. CAD, coronary artery disease. SNP, single nucleotide polymorphism. Reproduced by kind permission of UK Biobank ©.

### Phenotyping of clinical and demographic variables

CAD was defined as a history of coronary artery bypass grafting, myocardial infarction (MI), coronary artery angioplasty, or billing codes (OPCS-4: K40, K41, K45, K49, K50.2, K75) as previously performed.^16^ Prevalent and incident CAD status were defined by whether participants were diagnosed with CAD before or after the time of their MRI. Demographic variables were all ascertained at enrollment and included age, race, and sex of participants.

### Genotyping and genome-wide association study

The genotyping procedures of the UK Biobank have been described previously in detail.^15^ The genotyping arrays were the UK BiLEVE Axiom Array or the UK Biobank Axiom Array (both Affymetrix). The array-derived genotypes were imputed using the Haplotype Reference Consortium, UK10K, and 1000 Genome reference panels. Variant quality control measures included the following filters: MAF ≥ 1%, single nucleotide variant missingness <10% and HWE P ≥ 10−15, MAC ≥ 50, and INFO score ≥0.6. Sample quality control measures included excluding individuals if the single nucleotide variant missingness was equal to or exceeded 10%. Association analysis was performed in participants of European ancestry using REGENIE with adjustment for age, sex, and first ten PC of genetic ancestry.

### Extraction of splenic features

Briefly, the UK Biobank abdominal MRI protocol was as follows.^17^ The study aimed to image 100,000 healthy UK participants aged between 40 and 69 years old. 1.5 T clinical MRI scanners were utilized (Magnetom Aera, Siemens Healthineers, Erlangen, Germany) to acquire whole-body T1-weighted dual echo gradient echo (GRE) sequences. The parameters were as follows: echo times (2.39/4.77 ms), pixel size (2.23 × 2.23 mm2), slice thickness (3–4.5 mm), repetition time (6.69 ms), and flip angle (10°). For each patient, four MRI contrasts were available: in-phase (IP), out-of-phase (OP), water, and fat. We downloaded all abdominal MRIs from the UK Biobank.

We then used deep learning to segment spleens from abdominal MRIs of our study population and extracted 107 splenic radiomic features. Briefly, we used a stitching algorithm to stitch together MRI scans from six acquisition stations and compose whole-body scans outputted as four phases: water, fat, in phase, and out of phase (https://github.com/biomedia-mira/stitching).^34^ Utilizing a pre-trained nnuNet segmentation model, originally trained on 10,000 UK Biobank abdominal MRIs, we generated predictions of voxels corresponding to the spleen (code: https://github.com/BioMedIA/UKBB-GNC-Abdominal-Segmentation, trained models: https://gitlab.com/turkaykart/ukbb-gnc-abdominal-segmentation).^18^ This model had no errors in over 95% of the spleen segmentations in the UK Biobank data, and we performed no additional training. The models utilize a nnU-net architecture, a variant of the popular U-Net architecture that was shown to outperform U-Net on a range of biomedical imaging segmentation tasks. The models were validated in a previous study using 400 previously labeled images.^18^ The inputs to the model were water, fat, in- and opposed-phase stitched MRss. The model was applied on a Google Cloud Platform with CUDA version 11.6 and with 2 Tesla T4 GPUs available with 16 GB RAM each. Lastly, we extracted the voxels that corresponded to the spleen segment.

We applied the *pyradiomics* software (version 3.1.0) to the voxels identified by the model as spleen segments to extract shape and texture-based features.^21^ Generation of these features includes first-order statistics describing the image region and computation of the relationships between neighboring pixels. All code was parallelized using multi-processing to decrease runtime. In addition to the features extracted through this approach, we utilized the splenic volume features provided by the UK Biobank, which was determined using a deep learning U-net architecture as described in this study.^22^

### Correlation of splenic features with each other and cardiometabolic outcomes

We examined the associations of splenic features with age, sex, and BMI (https://biobank.ctsu.ox.ac.uk/showcase/field.cgi?id=21001). We used a linear regression model with each splenic feature as the independent variable and age at enrollment, sex, BMI, and days between enrollment and MRI acquisition as dependent variables. All splenic radiomic features were normalized to a distribution with mean 0 and standard deviation 1 for all analyses. We reported the coefficients and standard errors of both BMI and sex for each splenic feature.

We also associated the splenic features with blood-based biomarkers available in the UK Biobank. Blood-based markers include counts and percentages of basophils, eosinophils, lymphocytes, monocytes, neutrophils, platelets, reticulocytes, high light scatter reticulocytes, white blood cells, red blood cells, and nucleated red blood cells. Other biomarkers were C reactive protein, hematocrit, hemoglobin concentration, immature reticulocyte fraction, mean corpuscular hemoglobin, mean corpuscular hemoglobin concentration, mean corpuscular, platelet, reticulocyte, and sphered cell volumes, and platelet and erythrocyte distribution width (https://biobank.ndph.ox.ac.uk/ukb/label.cgi?id=9081). For each of the blood-based biomarkers, we implemented a linear regression model with each splenic feature as the outcome and the biomarker as a covariate and adjusted for age, sex, BMI, and the days between enrollment and the MRI acquisition. We then reported the coefficient, which can be interpreted as the change in one unit of the biomarker per 1 SD of the radiomic feature, and standard error of the biomarker in the model.

### Identification of splenic features associated with CAD

We examined for splenic radiomic features that are associated with CAD outcomes. We differentiate between CAD diagnosed prior to MRI (prevalent cases) for assessing splenic markers of existing CAD, and first CAD after MRI (incident cases among those without prevalent CAD) for assessing splenic predictors of future CAD. We performed feature processing before training two models for the outcomes of prevalent and incident CAD. Race and sex were coded as binary indicator variables. For each feature, we imputed any missing values with the median of all values for the feature, since missingness was less than 10%. We then employed forward selection to identify independent features for each CAD outcome, thereby minimizing potential collinearity. Starting with all features including splenic features, age, race, and sex, this method selected features one at a time that had a P value of less than a threshold when added to a model with already included features (https://github.com/AakkashVijayakumar/stepwise-regression/tree/master). We selected this threshold using 5-fold cross-validation on a held-out validation set, and our threshold options were 0.025, 0.05, 0.1, and 0.2. After a subset of features was selected, we standardized all features to normal distributions.

Subsequently, we analyzed the associations between the selected radiomic features and CAD outcomes using L1-regularized multivariable regression models, specifically logistic regression and Cox proportional hazards for prevalent and incident CAD respectively. For each model, 70% and 30% of the data were utilized for training and evaluation respectively. To identify splenic features associated with prevalent CAD, we trained an L1-regularized logistic regression model for the outcome of prevalent CAD. We optimized the logistic regression model using a 5-fold cross-validation grid search for various hyperparameters, including different regularization parameters (C = [5✕10^-5^, 5✕10^-4^, 5✕10^-3^, 0.05, 0.5, 1, 5, 10]), maximum number of training iterations (max_iter = [1000, 5000]), and reweighting of data points to minimize class imbalance (class_weight = [balanced, None]). We computed AUROC to evaluate the logistic regression. For the outcome of incident CAD, we used a Cox proportional hazards model in order to account for the temporal information of time from MRI acquisition to CAD diagnosis.

The time event was the days from MRI date to CAD diagnosis, and patients with CAD diagnosis before MRI date were excluded from the analysis. We computed concordance and AIC to evaluate the model. To ascertain the robustness of our findings, we performed 1000 resamplings using Monte Carlo bootstrapping on the test set to calculate 95% CI of the AUROC or concordance index.

### Genome-wide association study and gene prioritization

We explored the genetic underpinnings of CAD-associated splenic features by conducting GWAS on common variants (minor allele frequency > 0.01) for the fourteen splenic radiomic features. We used the PLINK (version 2.0) and REGENIE (version 3.2.8) software to run a GWAS for each splenic feature for chromosomes 1-22. We used a minor allele frequency of 0.01, missingness upper threshold of 0.1, and Hardy-Weinberg equilibrium value of 1*10^-15^. We adjusted for age, sex, first ten genetic PCs, and genotyping array. For all phenotypes, we computed the genomic inflation factor and the LD score intercept using LD Score Regression (LDSC) using LD scores from participants of European ancestry from the hapmap3 variants.^23^

To further analyze the results, we used the Functional Mapping and Annotation of Genome-Wide Studies (FUMA), a platform for annotation of GWAS results and gene prioritization.^24^ Independent, significant loci were detected based on a significance threshold of p < 5*10^-8^ and clumping with 1000 Genomes data, with an R^2^ threshold of 0.6. Lead SNPs were then detected based on clumping on independent, significant loci with an R^2^ threshold of 0.1. We used an online list comparator to identify overlapping lead SNPs (https://molbiotools.com/listcompare.php). For gene prioritization, we used FUMA to identify the nearest genes to each SNP and the genes prioritized by expression quantitative trait loci (eQTL).^24^ The nearest gene to each SNP was identified using a window of 10 Kb of the SNP. We combined the PoPS analysis with positional mapping in order to prioritize genes, as combining similarity-based and locus-based approaches has been shown to lead to better identification of causal genes.^25^ To implement PoPS, we first computed MAGMA scores from the summary-level results of the GWAS with each splenic feature. We then computed a PoPS score for all genes within 10 Kb of the significant SNPs. We selected the gene with the highest PoPS score in each locus. All GTEx v7 eQTL data were used for eQTL mapping, specifically adipose tissue, adrenal gland, blood, blood vessel, brain, breast, colon, esophagus, heart, liver, lung, muscle, nerve, ovary, pancreas, pituitary, pancreas, salivary gland, skin, small intestine, spleen, stomach, testis, thyroid, uterus, and vagina tissues. In order to prioritize genes using PoPS, we processed publicly available features derived from gene expression data from various organs (https://github.com/FinucaneLab/gene_features). For the GWAS results for each splenic phenotype, we then applied MAGMA, which provides gene-level association statistics. Finally, we applied the PoPS algorithm to derive scores for each gene.^26^ We stratified the genes by genomic locus and prioritized the gene with the highest PoPS score. For each splenic phenotype, we filtered genes prioritized by at least two of the three methods. We then compiled all genes prioritized in this manner for any of the ten splenic phenotypes.

From the genes prioritized for the splenic phenotypes, we used OpenTargets to identify genes associated with CAD. Associations with CAD are based on a combination of scores based on data from Open Targets Genetics, ClinVar, an NIH public archive of the relationship between human genetic variants and phenotypes, and other genetic sources (https://platform-docs.opentargets.org/evidence#open-targets-genetics). We included all genes as associated with CAD if the overall association was greater than 0. For the genes with non-zero associations with CAD, we then searched for the mouse phenotypes in mice where the gene was knocked out using the International Mouse Phenotyping Consortium, a collaboration between 21 research institutions where approximately 20,000 genes are systemically knocked out one by one in mice to understand the resulting phenotypes.^27,28^

### Overlap of SNPs and genetic correlation between splenic phenotypes and CAD

We used GWAS results from a previous meta analysis for CAD for determining overlap and to identify genetic correlation.^29^ We identified SNPs that were significantly associated with both CAD and at least one of the six splenic phenotypes. We used a p-value threshold of <5✕10^-8^ to define significant SNPs for both the CAD and splenic phenotype GWAS results. For each splenic phenotype, we clumped the significant SNPs overlapping with CAD using 1000 Genomes reference panel of European participants to identify lead SNPs.^23,30^ After filtering to SNPs meeting the genome-wide significance threshold, clumping of SNPs was performed using the default settings of 0.0001 as the significance threshold for index SNPs, 0.01 as the threshold for clumped SNPs, 0.50 as the LD threshold, 250 kb as the distance threshold, and 1000 Genomes patient cohort as the reference population. Next, we investigated the phenotype associations of the lead SNPs using PhenoScanner, a database that contains over 65 billion phenotype associations and 150 million unique variants.^31,32^ To compute genetic correlation, we used existing heritability estimation software and 1000 Genomes European LD score data.^23,30,33^

## Results

### Study population

Our study included 42,059 participants in the UK Biobank study who had abdominal MRIs without known hematological cancer at the time of MRI (**Supplemental** Figure 1). The study population at enrollment had a mean age of 55.1 years (standard deviation [SD] 7.5), body-mass index (BMI) of 26.1 kg/m^2^ (SD 4.2), comprised 52.1% females (N=21,895), and was predominantly of British White ancestry by self-report (96.7%, N=40,675). At MRI ascertainment, the prevalence of CAD, hypertension, hyperlipidemia, and type 2 diabetes was 4.7% (N=1,987), 24.0% (N=10,082), 16.7% (N=7,010), and 3.0% (N=1,243), respectively. The median time from UK Biobank enrollment to MRI was 9.4 years [IQR: 6.8-12.0], and the median follow-up time after MRI was 5.00 years [IQR: 3.85-6.63]. Key hematologic parameters measured at enrollment showed a mean white blood cell count of 6.6✕10^9^ cells/L (SD: 1.6), hemoglobin concentration of 14.2 g/dL (SD: 1.2), platelet count of 249.9✕10^9^ cells/L (SD: 56.3), and hsCRP levels at 2.1 mg/L (SD: 3.6) (**Table 1**).

**Table 1.**
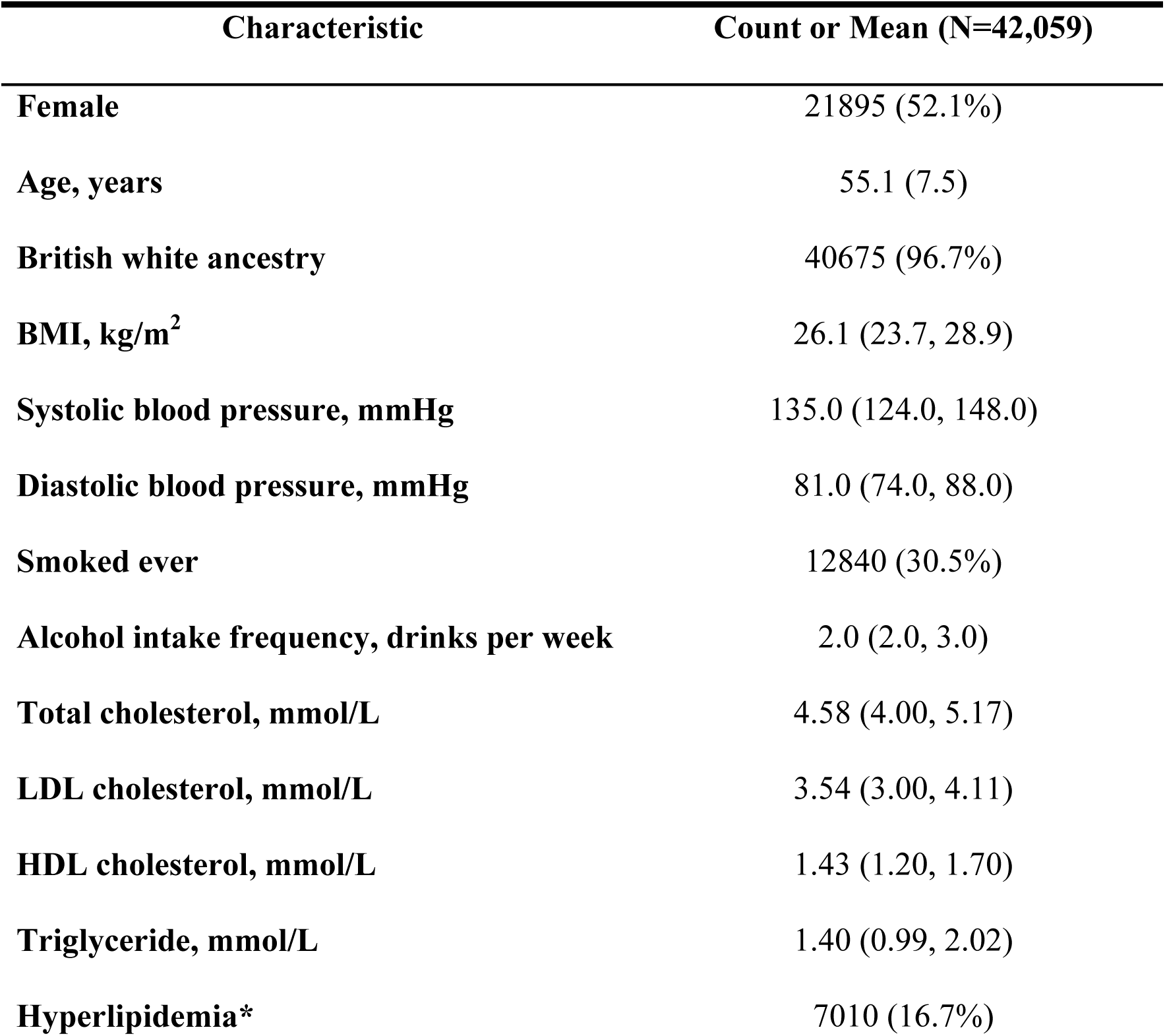
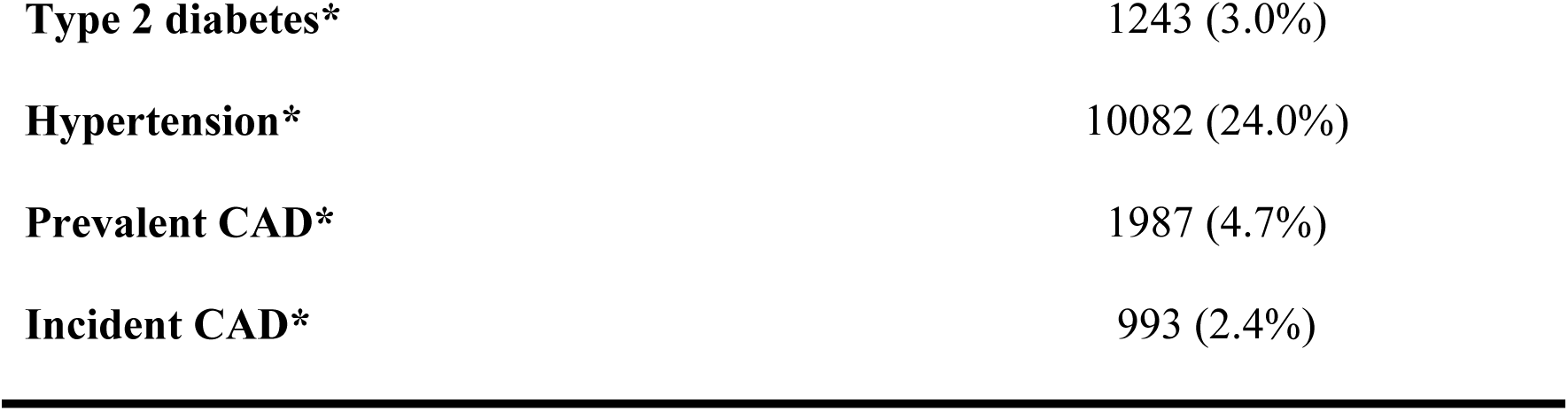
Baseline characteristics. The number of incident and prevalent CAD cases in the cohort is shown below, and the gender and race breakdown of the population is also presented. For binary variables, metrics are represented as n (%). For continuous variables, age is represented as mean (SD), and all other continuous metrics are reported as median (IQR). All variables are measured at enrollment, unless an asterisk is included, indicating measurement at MRI date or, in the case of incident CAD, measurement at any point after MRI date. CAD, coronary artery disease. BMI, body-mass index. LDL, low-density lipoprotein. HDL, high-density lipoprotein. CAD, coronary artery disease. BMI, body mass index. LDL, low-density lipoprotein. HDL, high-density lipoprotein. *measured at MRI date.

### Deep learning-extracted radiomic characteristics of the spleen

In our study population, splenic volume was previously annotated by the UK Biobank centrally for 15,215 participants with a mean of 0.17 liters (SD 0.07). Splenic volume varied with age and sex. It decreased modestly with age in this middle-aged cohort, from 0.18 mg/g (SD: 0.07) among individuals aged 40-48 years to 0.16 mg/g (SD: 0.07) among those aged 62-70 years. Splenic volumes on average were lower in women (mean 0.14 mg/g, SD 0.05) compared to men (mean 0.19 mg/g, SD 0.07).

We generated spleen images from the first MRI for all 42,059 participants. We extracted 107 radiomic features using the pyradiomics software (version 3.0.1).^21^ Features are grouped into first order statistics, 3D shape-based features, and five categories of gray level information (Figure 2 and **Supplemental Table 1**).

**Figure 2.**
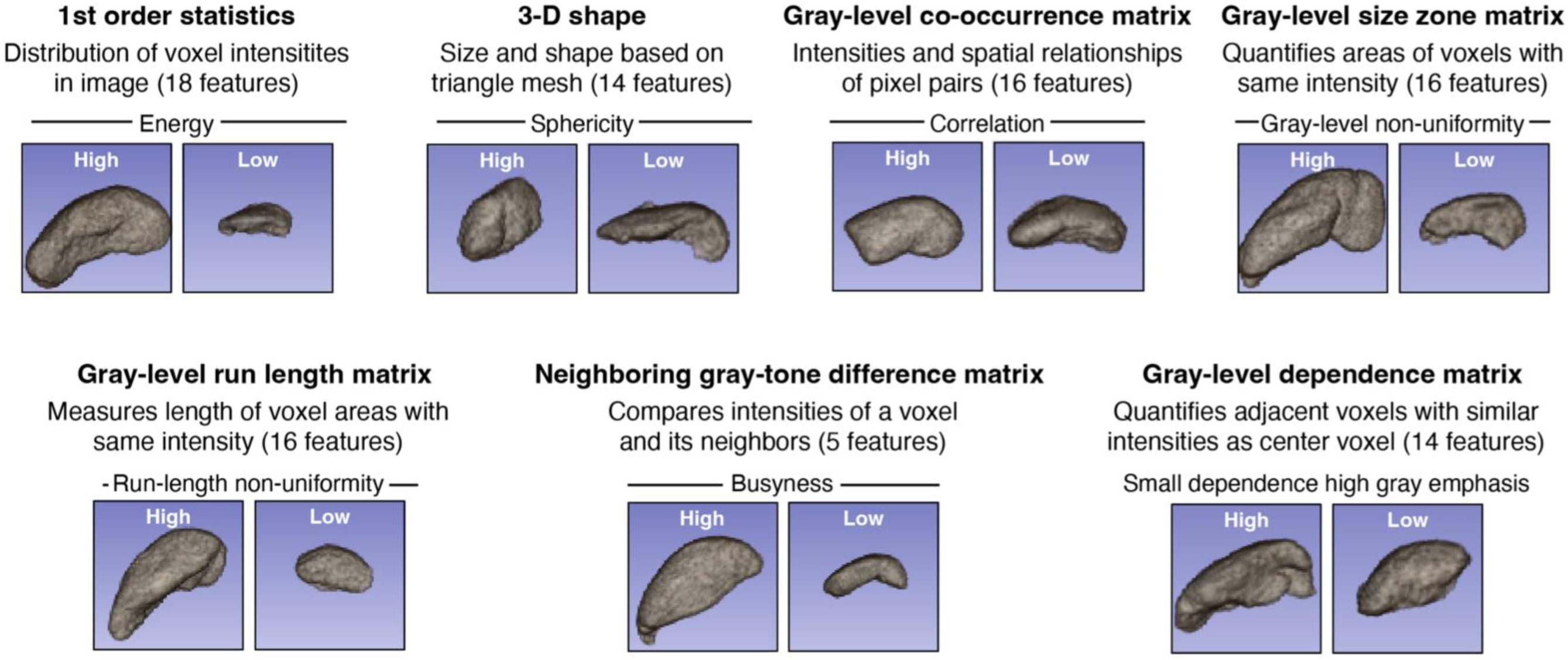
Categorization of Extracted Splenic Radiomics Features. 7 categories of radiomic features with descriptions, numbers of quantified features, and visualizations of high and low values for a selected feature. Reproduced by kind permission of UK Biobank ©.

We extracted 18 first-order statistics that indicate the distribution of voxel intensities within the masks of the image region. These features capture the magnitude, randomness, uniformity, and asymmetry of the voxel values, as well as standard descriptors such as mean, median, and range. We derived 14 shape-based 3D metrics gleaned from the approximated shape defined by the triangle mesh independent of gray-level intensities using a ‘marching cubes’ algorithm.^35^ These features are readily interpretable. As expected, several volume-related features, including mesh volume, voxel volume, major and minor axis lengths, and surface area are highly correlated with the annotated volume which was measured by the UK Biobank as part of the imaging exam (Pearson correlation coefficients [ρ] ranging from 0.70 to 0.99; all P<0.001). In contrast, morphologic measures such as sphericity, elongation, and flatness exhibited relatively lower or no correlation with the annotated volume (ρ < 0.25), indicating their orthogonal informational value (**Supplemental Table 2**).

The remaining 75 features focused on texture metrics relating to gray levels. We extracted gray level co-occurrence matrix (GLCM) to measure pixel intensity pairings within a spatial context, the gray level size zone matrix (GLSZM) to count interconnected voxels zones of similar grayness, and the gray level run length matrix (GLRLM) to assess the spatial distributions of these zones, reflecting graininess. 16 features were generated using each of these matrices. The neighboring gray tone difference matrix (NGTDM) estimates the variations in gray value over a specified distance for 5 features, and the gray level dependence matrix (GLDM) gauges the connectivity of voxels relative to a center voxel across 14 features. **Supplemental Figure 2** shows Pearson correlation coefficients between features, and further details are in https://pyradiomics.readthedocs.io/en/latest/features.html and **Supplemental Table 1.**

### Splenic radiomics with other variables

Given the known influences of age, sex, and obesity on splenic function, we examined the association of age, sex, and BMI (after adjustment for the others) with each splenic feature using multivariable linear regression and observed many significant associations. In particular, sex showed the strongest associations with splenic size including minor axis length (0.7 SD lower in females vs males, 95% CI [0.68,0.72]) and surface area (0.70 [0.68,0.72]). BMI was most significantly associated with several texture features: one unit increase in BMI was associated with 0.11 [95% CI: 0.10, 0.11], 0.09 [0.09, 0.10], 0.09 [0.09, 0.09] SD increase in GLSZM gray level non-uniformity, run length non-uniformity, and GLRLM gray level non-uniformity, respectively (Figure 3 and **Supplemental Figure 3**).

**Figure 3.**
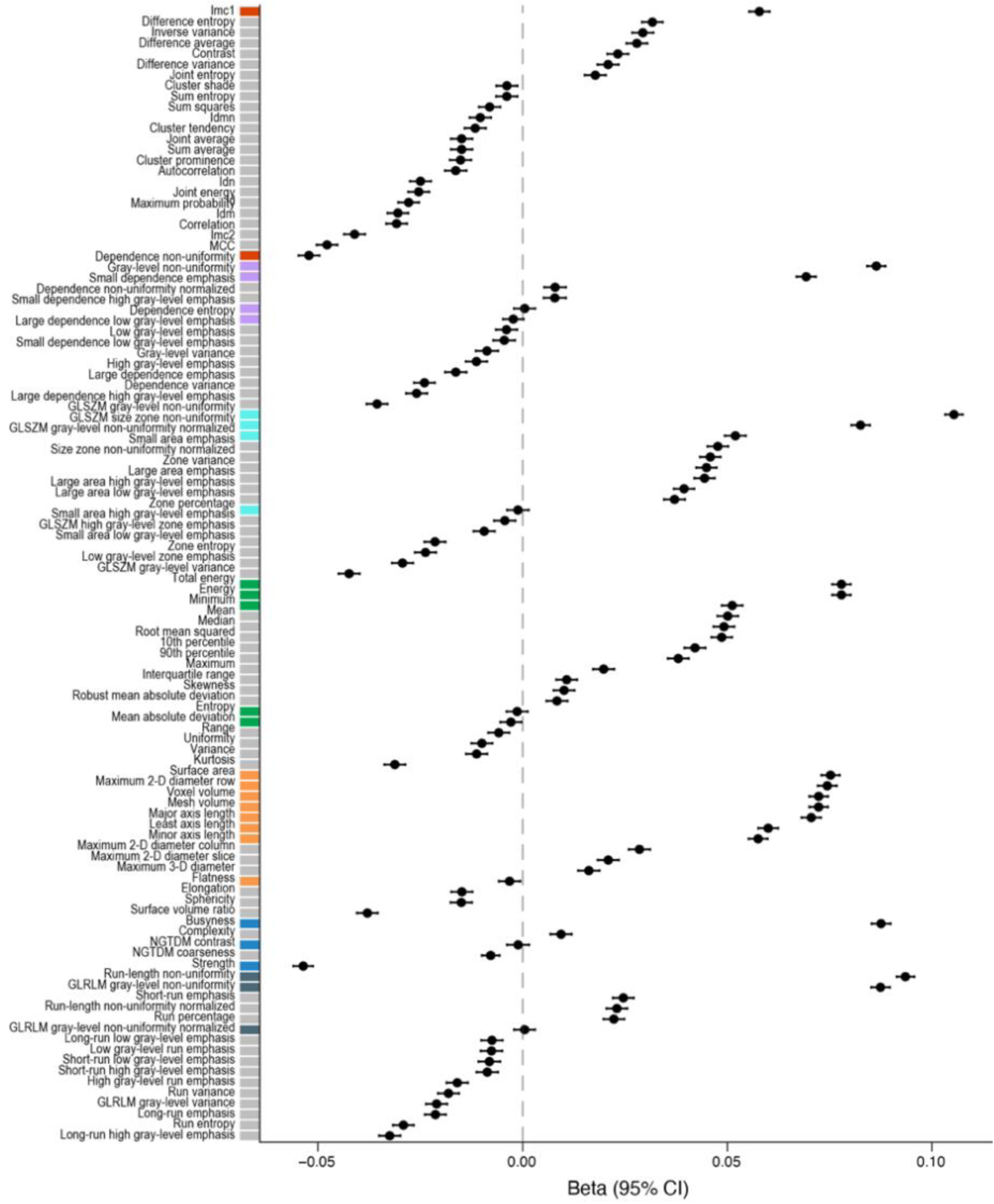
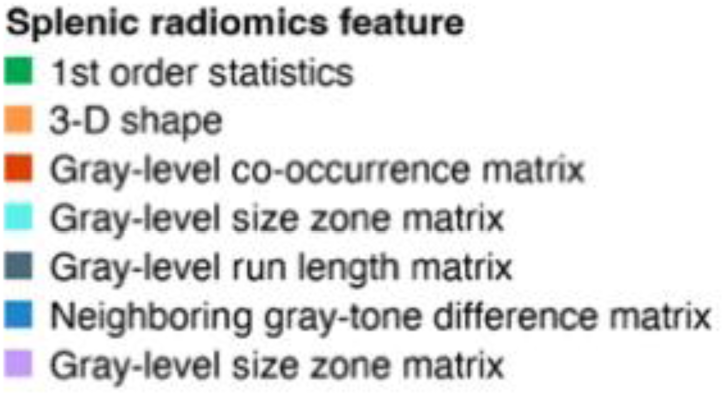
Coefficients of BMI in a linear regression model for each splenic feature, adjusting for age and sex. Splenic radiomic features are grouped and colored by category using the color scheme from Figure 2. Features are grayed out if the Bonferroni corrected p-value for the coefficient of the BMI feature is greater than or equal to 0.05.

We then examined the associations between splenic features and hematologic biomarkers, adjusting for age, sex, and BMI. The strongest associations were of energy and GLSZM size zone non-uniformity exhibited with high light scatter reticulocyte count, with an increase of 0.21 [95% CI: 0.21, 0.21] per 1 SD of each radiomics feature. Many splenic features were negatively associated with mean spherical cell volume, including surface area (**Supplemental Figure 4**). The strongest association for white blood cell (WBC) count was a 0.10 [0.09, 0.11] increase for 1 SD increase of GLCM informational measure of correlation 1. For red blood cell count, a 0.16 [0.15, 0.17] increase was associated with 1 SD increase of GLSZM size zone non-uniformity.

For C-reactive protein, a 0.05 [0.03, 0.06] increase was associated with 1 SD increase of median. **Supplemental Table 3** contains the top splenic radiomic features associated with each hematological parameter.

### Prioritizing CAD-associated splenic radiomics

For prevalent CAD, the optimized regression model achieved an AUROC of 0.77 (95% CI 0.75-0.78) in the held-out test set (N=12755), and the Cox model for incident CAD yielded a concordance index of 0.68 (95% CI 0.65-0.71) in the test set (N=12022). Notably, 9 and 5 splenic radiomic features were retained in the prevalent and incident CAD models, respectively, achieving statistical significance (P<0.05) after adjustment for other covariates.^36^ There is no overlap in significant splenic features between prevalent and incident CAD. For prevalent CAD, associated features included GLSZM gray level non-uniformity (OR per 1 SD increase: 1.59 [95% CI: 1.38, 1.82], P<0.001, FDR<0.001) and sphericity (OR: 1.16 [95% CI: 1.09, 1.23], P<0.001, FDR<0.001), among others. GLCM correlation, energy, GLDM metrics of small dependence high gray level emphasis and gray level variance, GLSZM large area low gray level emphasis, GLRLM run length non-uniformity, and GLCM inverse difference also showed significant associations with prevalent CAD. For incident CAD, associated features included GLRLM run length non-uniformity (HR: 1.17 [95% CI: 1.09, 1.25], FDR<0.001), which was also associated with prevalent CAD, and GLCM inverse difference normalized (HR: 0.90 [95% CI: 0.85, 0.95], FDR<0.001) (**Supplemental Table 4,** Figure 4A-B). All features significantly associated with prevalent or incident CAD met the FDR threshold of 0.05 for significance for genetic discovery and were used for subsequent analyses.

**Figure 4.**
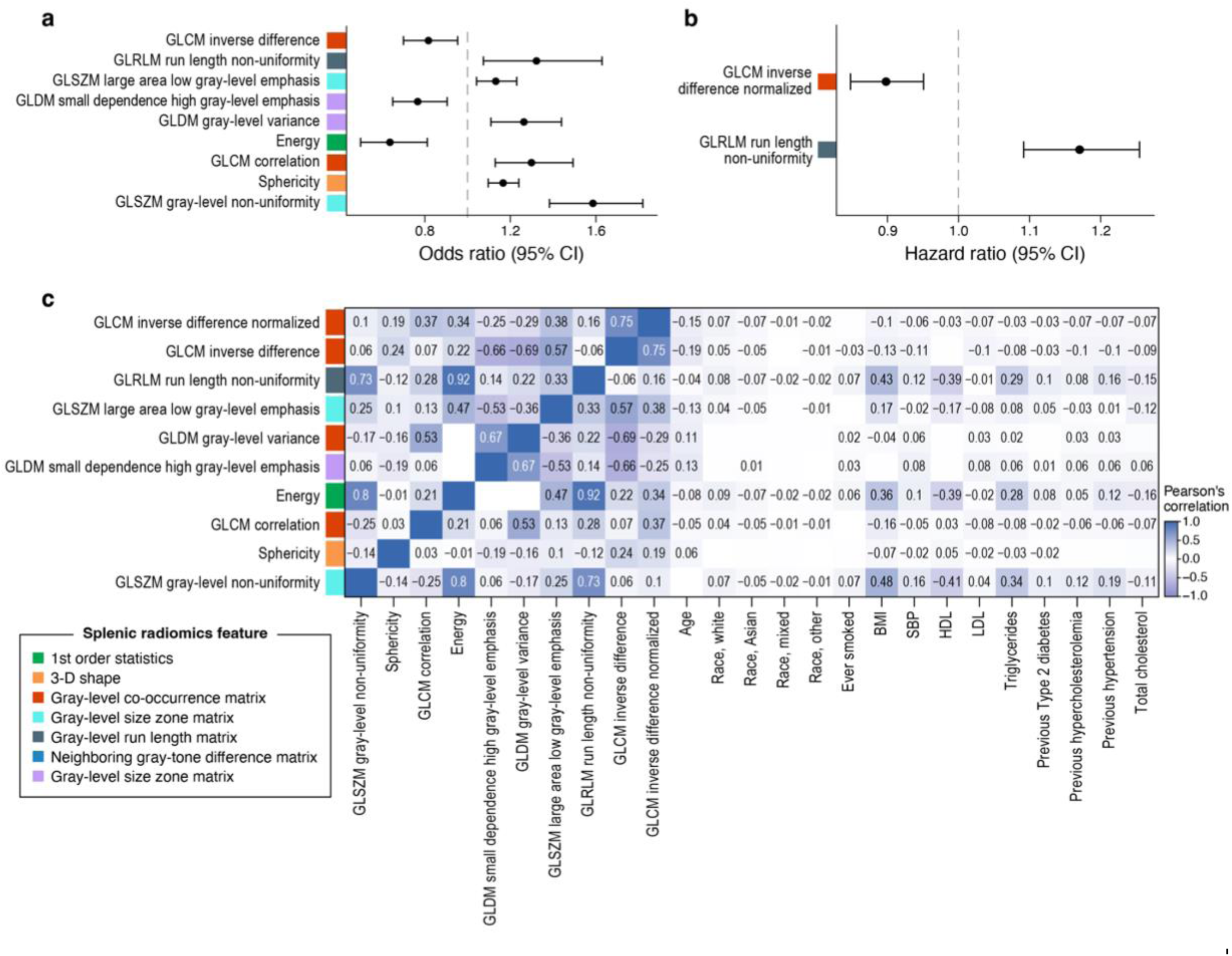
Splenic radiomics features selected by prediction models for prevalent and incident CAD. a) Splenic radiomics features that were nominally associated (p-value < 0.05) with the prevalent CAD in a logistic regression model. b) Splenic radiomic features that were also nominally associated with incident CAD in a Cox regression survival analysis model among those without prevalent CAD. Covariates for both models included age, race, sex, and a set of splenic features chosen by forward stepwise regression, to ensure that no splenic features in the model were significantly correlated with each other. c) Correlations of the fourteen nominally significant splenic features across both models with conventional CAD risk factors. The Pearson correlation coefficient is shown for correlations that are significant. Higher gray non-uniform, gray level variance, and run-length non-uniformities values are associated with increased texture variation. Correlation refers to the correlation between voxel locations and their gray level intensities. A higher energy value indicates higher gray intensities. Higher small dependence high gray level emphasis indicates increased variation of areas of high gray levels. Higher large low gray level emphasis indicate decreased variation of areas of low gray levels respectively. Higher inverse difference reflects decreased texture variation of the spleen. Feature abbreviations are as follows: Coronary Artery Disease, CAD. Energy, firstorder_Energy. Sphericity, shape_Sphericity.

To examine the relationships between these CAD-associated splenic features and conventional CAD risk factors, including age, sex, race, smoking, BMI, diabetes, hypertension, and total, HDL, and LDL cholesterol levels, we calculated their pairwise Pearson correlations. Gray level-uniformity, energy, and run-length non-uniformity are moderately positively correlated with BMI and triglyceride levels, and all three features negatively correlate with HDL cholesterol. Overall, most features exhibit only weak correlations with all conventional CAD risk factors (Figure 4C). **Supplemental Figure 5** shows representative MRI images for the prioritized splenic features.

### 219genome-wide significant regions associated with CAD-associated splenic features

In the GWAS for the fourteen splenic radiomics features, there was no significant inflation of association statistics (*λ*_GC_ ranges from 1.03 to 1.15; LD score intercept ranges from 1.03 to 1.17. **Supplemental Table 5**). The genetic signals varied across the 14 traits. Using P < 5*10^-8^ and r^2^ < 0.1 as thresholds to identify significant and independent variants, we discovered 95 independent significant SNPs for sphericity, 72 for energy, 41 for GLRLM run length non-uniformity, 21 for GLSZM gray level non-uniformity, and 16, 9, 7, 4, 2, and 0 for GLSZM large area low gray level emphasis, GLCM inverse difference, GLCM inverse difference normalized, GLCM correlation, GLDM small dependence high gray level emphasis, and GLDM gray level variance, respectively. At the locus level, chr9:91392686, chr12:112037450, chr12:112007756, and 12:113165247 were all associated with 4 splenic features respectively; a few other discovered loci also associated with more than one feature, but more were associated with unique traits (**Figure 5, Supplemental Figures 6-14**).

**Figure 5.**
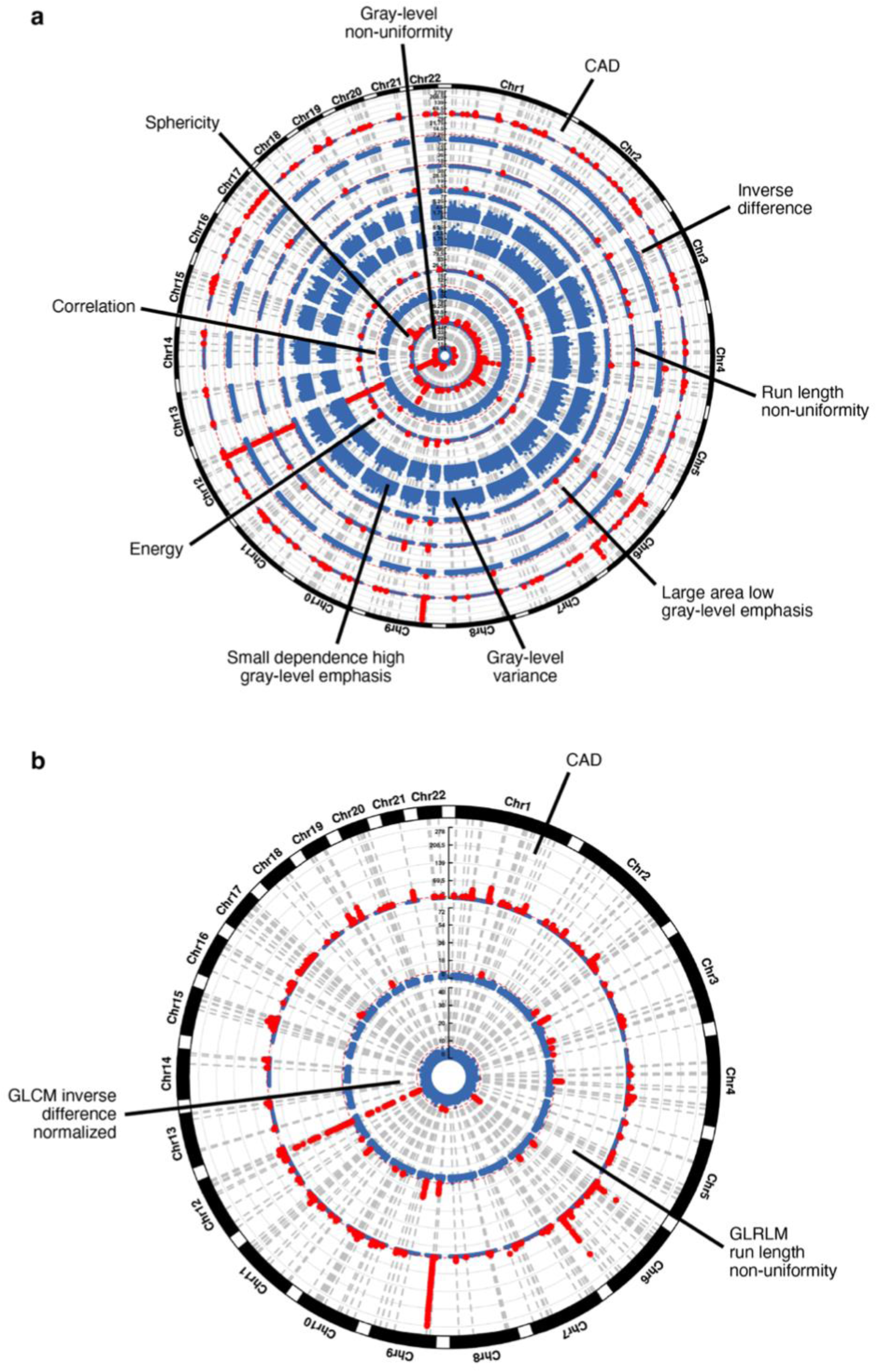
Circular Manhattan plots from GWAS with 14 splenic phenotypes and CAD. a) The circular Manhattan plot portrays the features that were statistically significant for prevalent CAD. From outside to inside, the features are CAD, GLCM inverse difference, GLRLM run length non-uniformity, GLSZM large area low gray level emphasis, GLDM gray level variance, GLDM small dependence high gray level emphasis, energy, GLCM correlation, sphericity, and GLSZM gray level-non uniformity. Red dots indicate significant loci. The y-axis is the log10 of the p-value. b) The circular plot shows the features statistically significant for incident CAD. From outside to inside, the features are CAD, GLRLM run length non-uniformity, and GLCM inverse difference normalized. Feature abbreviations are as follows: CAD, Coronary Artery Disease. Correlation, glcm_Correlation. Energy, firstorder_Energy. Sphericity, shape_Sphericity.

Utilizing GWAS results of CAD-associated splenic features, we assessed their genetic correlations with CAD, observing varying degrees of correlations. The features with the strongest correlations that had the same direction of effect on CAD as in the regression models were GLCM correlation (r_g_=0.17, P=0.002) and energy (r_g_=-0.12, P=0.01), indicating shared genetic basis with CAD. A few features had more modest genetic correlations with CAD, suggesting the need for studying the non-genetic pathways linking them with CAD (**Supplemental Table 15)**.

### THBS1, PDE5A, and 35 more CAD-associated genes are likely to be causal genes for splenic features

For GWAS of each CAD-associated splenic feature, we prioritized genes likely to be causal using three methods: 1) gene annotation based on distance (i.e., nearest gene), 2) polygenic priority score (PoPS), and 3) eQTL mapping based on cis-eQTLs. These loci mapped to 83, 58, 35, 21, 16, 9, 7, 4, 2, 0 respective genes based on proximity, by choosing the closest gene to each SNP within 10 Kb, for the splenic phenotypes listed in the order from the previous section (**Supplemental Tables 6-14**). The strongest signals for sphericity and GLDM small dependence high gray level emphasis were annotated to *TLX1NB* and *LRRC37A2:ARL17A,* and the signals for the other features were near *ATXN2,* a multi-functional gene linked to circadian rhythm and neurodegenerative diseases and prioritized in a previous GWAS for splenic volume.^22,37^

Using PoPS, we prioritized 0 to 48 genes per feature, with top putative causal genes including *S1PR3*, *ARHGAP42*, *SMG6*, ^38^*IRS1*, and *THBS1*, which were prioritized for 3 or more splenic features. *S1PR3* encodes a lysophospholipid mediator that has been shown to have both protective effects against stroke and vasoconstrictor effects.^39^ We observed strong corroboration between prioritized genes by PoPS (similarity-based approach) and distance (locus-based), increasing confidence in the results (**Supplemental Tables 16-24**).^26^ Using eQTL data from GTEx v7,^40^ top genes prioritized by eQTL mapping prioritized include *S1PR3*, *EGF*, *HECTD4, ARHGAP42*, *NAA25*, and *SMG6*, which were all prioritized for at least 4 splenic features, and are similar to those prioritized by nearest genes and PoPS (**Supplemental Tables 16-24**). Collectively, 119 genes were prioritized by at least two gene prioritization methods across all phenotypes (**Supplemental Table 25** and **Figure 6**).

**Figure 6.**
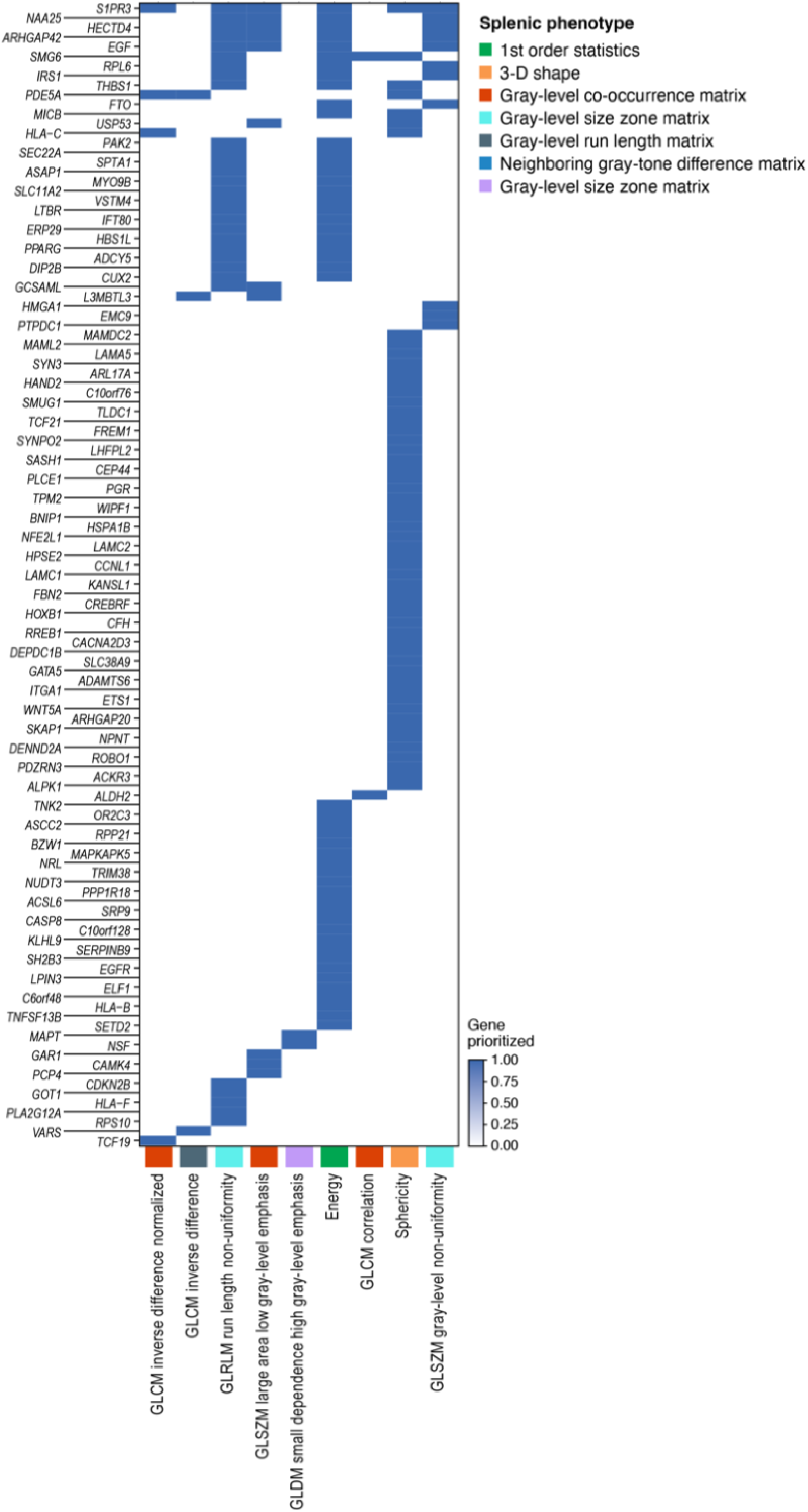
Genes prioritized for CAD-associated splenic radiomics. Genes were only included if they were prioritized by at least two methods (out of nearest gene, eQTL, PoPS) for at least one of the fourteen CAD-associated splenic phenotypes. Genes are grouped by the most associated splenic phenotypes from left to right. In addition, genes prioritized by similar phenotypes are grouped together. Feature abbreviations are as follows: Correlation, glcm_Correlation. Energy, firstorder_Energy. Sphericity, shape_Sphericity.

We explored the functional implications of genes prioritized for their links to CAD, leveraging OpenTargets to assess their CAD associations and the availability of targeted therapies. Among these, 37 genes, including *EGF, HECTD4, ARHGAP42, NAA25, SMG6, RPL6, IRS1, THBS1, PDE5A, FTTO, PPARG, CUX2,* have established CAD associations based on various genetic data sources (**Supplemental Table 25, Methods**).^31,32^ These genes are involved in multiple mechanisms, including inflammation (e.g., *THBS1*), smooth muscle cell regulation (e.g., *TCF21*, *PDE5A*), hypertension (e.g., *HECTD4, ARHGAP42*), heart tissue development (e.g., *WNT5A*, *HAND2*, *TCF21*), and adipogenesis (e.g., *FTO, PPARG*). We traced back the 37 genes to our GWAS across splenic features and found many were discovered from energy, run-length non-uniformity, and sphericity GWAS (**Supplemental Table 25**). For each gene, we identified mouse phenotypes resulting from gene knockout using the International Mouse Phenotyping Consortium.^27,28^ Knocking out *SMG6*, *PDE5A*, and *TCF21* resulted in abnormal spleen morphology, enlarged spleens for *SMG6* and *PDE5A,* and small spleens for *TCF21*. *TCF21* knockout led to abnormal blood vessels. *THBS1* knockout led to abnormal and enlarged hearts (**Supplemental Table 25**).

### Overlap of SNPs and genetic correlation shed light on the link between splenic phenotypes and CAD

Utilizing previously published CAD GWAS,^29^ we compiled SNPs associated with CAD and identified the ones associated with splenic features. 396 and 390 CAD-associated SNPs were associated with energy and run-length non-uniformity respectively, and the overall median [IQR] number of SNPs associated with CAD and the splenic features was 255.5 [18, 337]. After clumping of SNPs, 24 and 22 independent CAD-associated SNPs were significantly associated with energy and run-length non-uniformity, respectively. The overall median [IQR] number of SNPs associated with CAD and the splenic features was 9 [3, 17], with 39 unique ones across all splenic phenotypes (**Supplemental Table 26**). We filtered to 35 lead SNPs where the effect direction of the SNP on CAD was consistent with the effect of at least one radiomic feature on CAD risk.

We interrogated the existing associations of lead SNPs using PhenoScanner^31,32^ to assess for pleiotropic associations (**Supplemental Table 27)**. Of the 35 SNPs, 7 (20%) were not associated with any known cardiovascular risk factor, including hypertension, diabetes, systolic and diastolic blood pressure, smoking, total, HDL, and LDL cholesterol, triglycerides, or weight (**Supplemental Figure 15**). These SNPs were rs7036656 (chr9p21.3), rs56750693 (chr12q24.12), rs11515 (chr9p21.3), rs4239427 (chr18q11.2), rs4098854 (chr12q24.12), rs1208250 (chr6q23.2), and rs1208258 (chr6q23.2). These SNPs were associated with GLSZM gray non-uniformity, energy, GLSZM large area low gray level emphasis, GLRLM run length non-uniformity, GLCM inverse difference, sphericity, and GLDM large dependence high gray level emphasis.

The top SNPs at two identified loci, rs7036656 and rs11515, are at the chr9p21 locus, the most strongly associated CAD locus but previously with limited mechanistic insight.^41^ The rs7036656 SNP is significantly associated with energy (P=1.5✕10^-20^), GLRLM run length non-uniformity (P=1.6✕10^-16^), GLSZM large area low gray level emphasis (P=8.0✕10^-9^), GLSZM gray non-uniformity (P=3.4✕10^-9^), and GLCM inverse difference (P=3.2✕10^-8^). The rs11515 SNP is significantly associated with energy (P=2.4✕10^-11^) and run length non-uniformity (P=3.3✕10^-9^). Both loci are associated with energy and run-length non-uniformity. The strongest signal in the GWAS for both energy and run-length non-uniformity was at the same locus, rs653178 (energy: P = 1.3✕10^-106^, Z score = 21.9; run_length non-uniformity: P = 9.2*10^-72^, Z score = 17.9; nearest gene: *ATXN2*), indicating further genetic overlap between the two radiomics features. This locus is associated with systolic and diastolic blood pressure.^42^

## Discussion

In this study, we harnessed deep learning to extract splenic phenotypes not readily quantifiable through conventional methods, establishing the link between spleen and CAD. We discovered several radiomic features, such as heightened sphericity, increased texture variation, and reduced gray level intensity in the spleen, that were robustly associated with elevated CAD risk. We explored the genetic underpinnings of these CAD-associated splenic features, providing insight into the potential mechanism of the spleen’s involvement in key processes related to CAD, such as inflammation, smooth muscle cell regulation, and hypertension. Notably, we mapped seven genetic loci unlinked to known CAD risk factors to the splenic features, offering potential new targets for intervention and dissecting the splenic axis of CAD.

Our study has several implications. The first is that novel deep learning techniques to non-invasively extract radiomic features in the spleen at scale enable association study and genomic analysis of splenic variation in the population. This approach is particularly pertinent for the spleen, an organ with limited annotations even in clinical reports. Furthermore, in our study, the splenic radiomic features carry detailed information on shape, size, texture, and intensity much beyond known splenic markers - except for volume-related splenic features highly correlated with known splenic volume, other features provided orthogonal information about the spleen. Lastly, the pipeline we built offers a scalable framework for extracting features of other organs from imaging, facilitating the construction and testing of novel biomedical hypotheses.

Second, we put the computer-learned features in a disease context and identified potential radiomic markers for CAD. For example, image-derived texture variation has been used to identify specific patterns within lymphoma, splenic infarction, and splenic cysts^43^ ; specific to splenic features, sphericity and flatness have previously been used to distinguish between lymphoma subtypes.^14^ Our work expanded their use to look across all splenic radiomic features, capturing several aspects of spleen, and comprehensively examined the potential markers of CAD. We also identified splenic features common to patients both before and after CAD diagnosis, specifically run-length non-uniformity, suggesting that increases in splenic texture variation occur before CAD diagnosis and persist after diagnosis. This finding provides evidence that splenic changes are present with early development of CAD and are not simply effects of later disease progression.

Third, we integrated genetics and yielded important discoveries on the potential mechanism linking the spleen to CAD. Through GWAS and subsequent gene prioritization and annotation, we identified causal genes of CAD-associated splenic features and found their strong relevance in inflammation, smooth muscle cell regulation, and hypertension. For example, a top prioritized gene *THBS1* is implicated in angiogenesis and inflammation; *PDE5A*, essential for smooth muscle cell relaxation and linked to CAD through dysfunctional nitric oxide signaling and the second messenger cGMP in atherosclerosis, and *TCF21*, a regulator of coronary artery smooth muscle cell precursors, were prioritized.^44,45,46^ Both *PDE5A* and *TCF21* knockouts in mice affect gross spleen morphology, highlighting their relevance to both CAD and splenic phenotypes and thus the validity of our findings.

Also, we identified 35 pleiotropic loci associated with CAD and splenic features, where the effect of the locus on the radiomics feature and CAD was consistent. Among them, 7 were not linked to any conventional CAD risk factors, suggesting orthogonal information of the splenic axis of CAD; in particular, rs7036656 and rs11515 on the Chr9p21 locus, one of the strongest CAD loci whose mechanism remained unclear since its initial discovery in 2007, is identified in our study as associated with splenic texture changes, such as energy and run length non-uniformity.^47^ These findings, together, shed light on novel mechanisms linking the spleen to CAD, providing potential targets for therapeutic intervention to address this unexplored axis.

Our study has limitations. Firstly, the UK Biobank cohort includes participants of mostly European ancestry, and the participants were recruited between the ages of 40 and 59, limiting the generalizability of our findings to other ancestries and younger patients. These results should be replicated for a more diverse cohort. Second, we included participants whose MRI were categorized as “high-quality” by the segmentation model and filtered out “low-quality” ones where the spleen was not identified. However, those filtered images may contain unique information that resulted in the classification. Third, to increase discovery power, we used a more liberal CAD definition, and therefore some associated splenic features may not be directly relevant to the etiology of strictly defined CAD.

In conclusion, by extracting novel splenic radiomics features linked to CAD and uncovering their genetic underpinnings, our work examined the unaddressed splenic axis of CAD. We demonstrated significant associations of splenic sphericity and texture variation with CAD risk, alongside identifying genetic variants and prioritizing genes tied to these spleen-CAD links. Leveraging several databases, we explored the functions of these genes and demonstrated their relevance and potential mechanisms to CAD etiology. Notably, we highlighted several loci, such as Chr9p21, linked to both splenic alterations and CAD yet unassociated with conventional CAD risk factors, presenting them as potential novel targets for therapeutic intervention.

Together, our work presents a new framework to uncover the underexplored splenic axis of CAD.

## Data Availability

UK Biobank individual-level data are available for request by application (https://www.ukbiobank.ac.uk). All code used for the described analysis will be uploaded to GitHub once the manuscript is accepted for publication. All extracted splenic radiomics features will be submitted to UK Biobank once the manuscript is accepted for publication.

## Acknowledgments

We gratefully acknowledge the participants who provided biological samples and data for UK Biobank. We also thank Mrs. Leslie Gaffney from the Broad Research Communication Lab for her valuable assistance in improving the display items.

