## Supplemental Figures for "Deep learning-derived splenic radiomics, genomics, and coronary artery disease" (from another version of this preprint: version 1, 2024-08-17)

**Supplemental Figure 1.** Flow diagram of patient cohort selection.

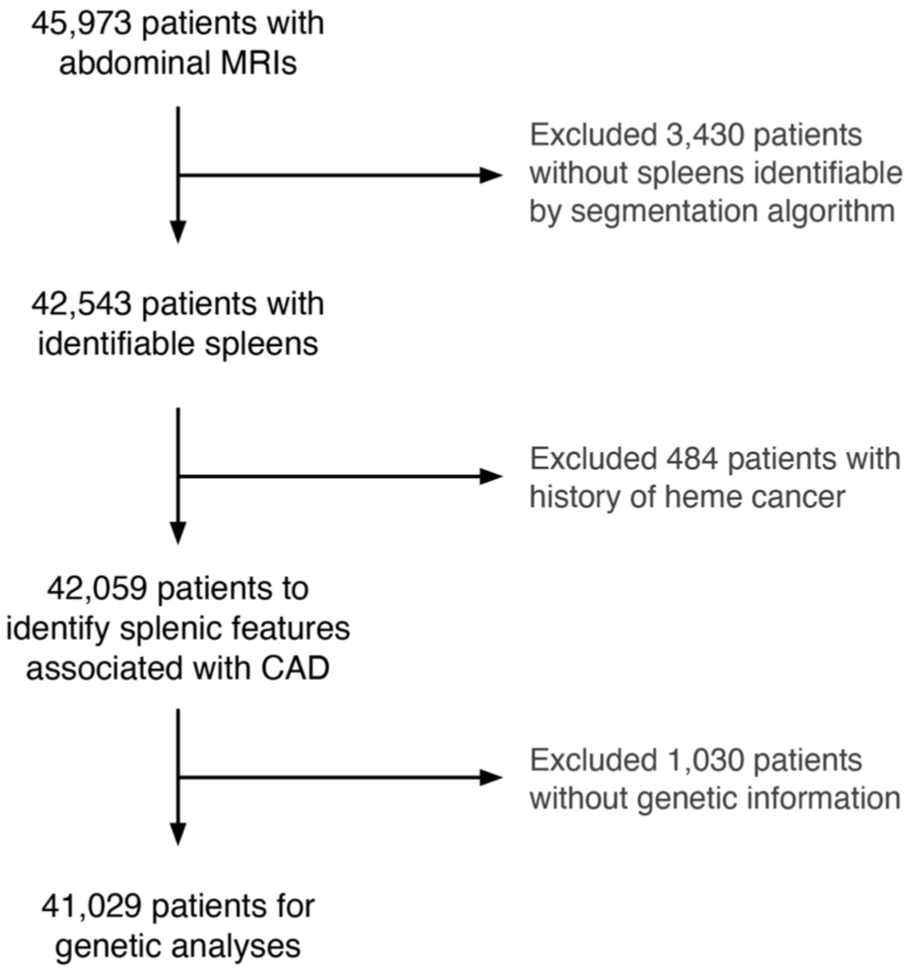

**Supplemental Figure 2.** Pearson correlations between splenic radiomic features. The heatmap showcases the pairwise Pearson correlations between the 107 imaging features extracted from spleens in 42,543 abdominal MRIs. Features in the shape category show very little correlation with other features.

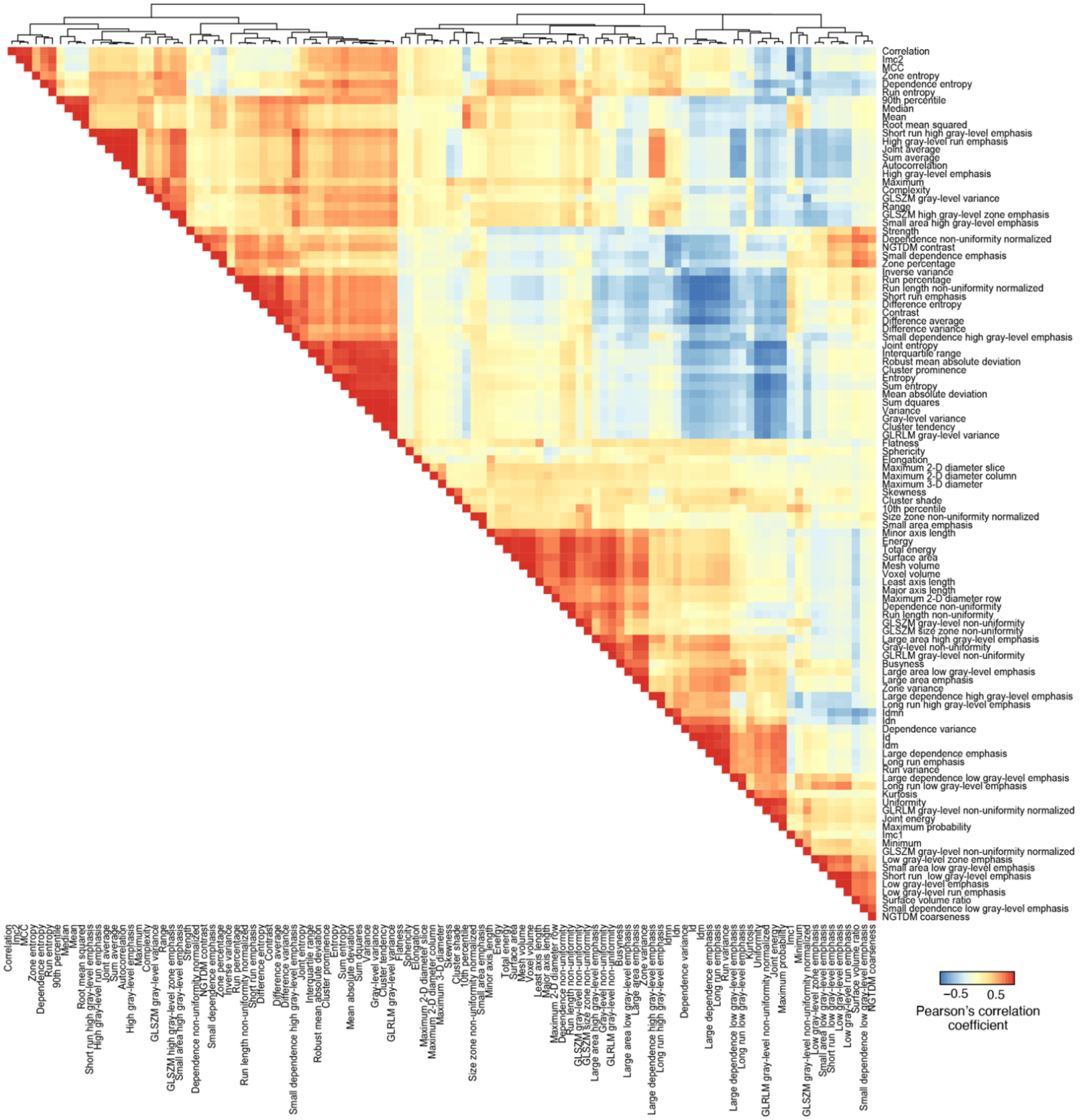

**Supplemental Figure 3.** Coefficients of sex in a linear regression model for each splenic feature, adjusting for age and BMI. Splenic radiomic features are grouped and colored by category. Features are grayed out if the Bonferroni corrected p-value for the coefficient of the sex feature is greater than or equal to 0.05.

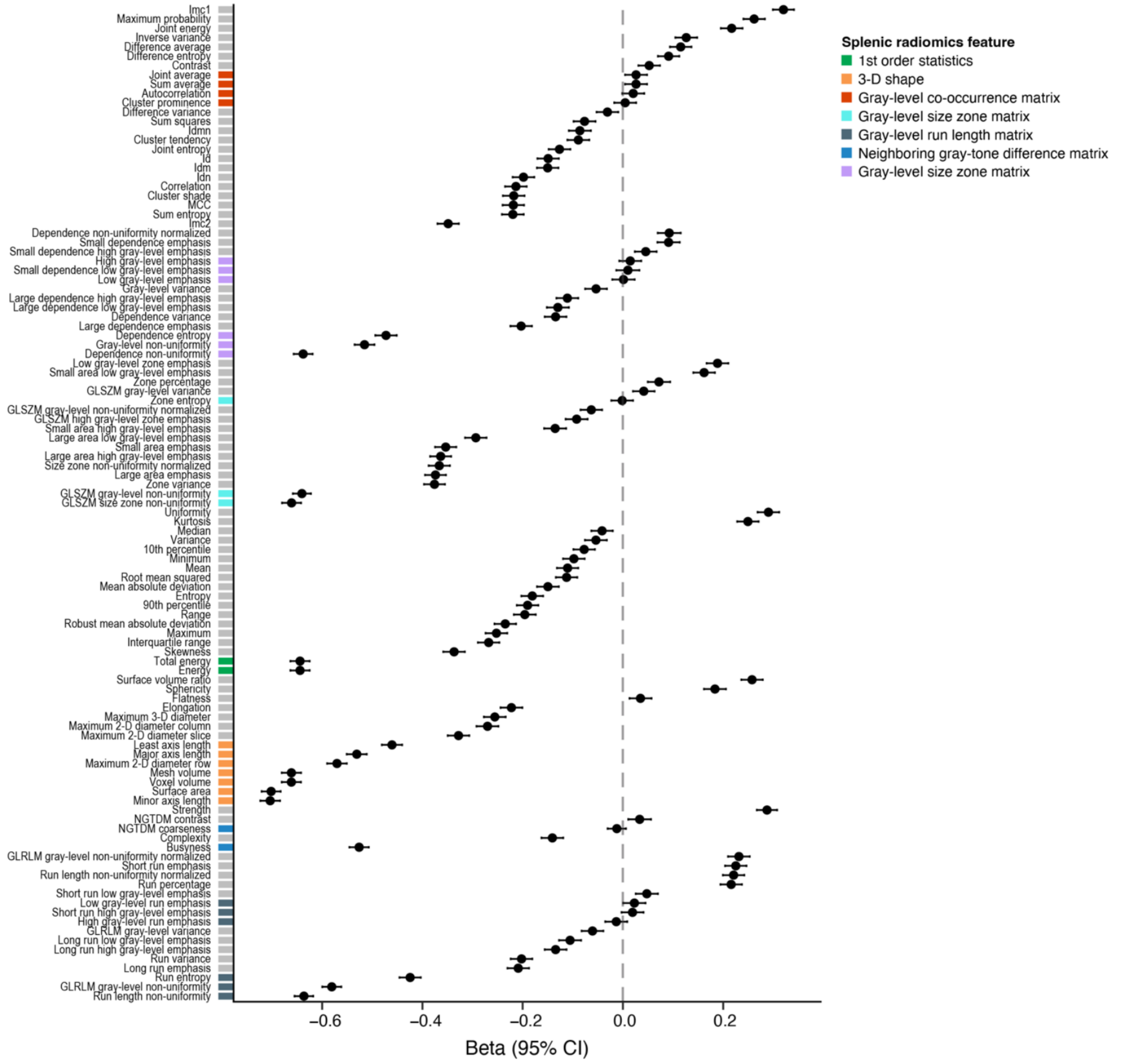

**Supplemental Figure 4.** Associations between splenic features and hematological biomarkers. The heatmap shows the Z-scores (coefficient divided by the standard error) for each hematological marker in the model for each splenic feature.

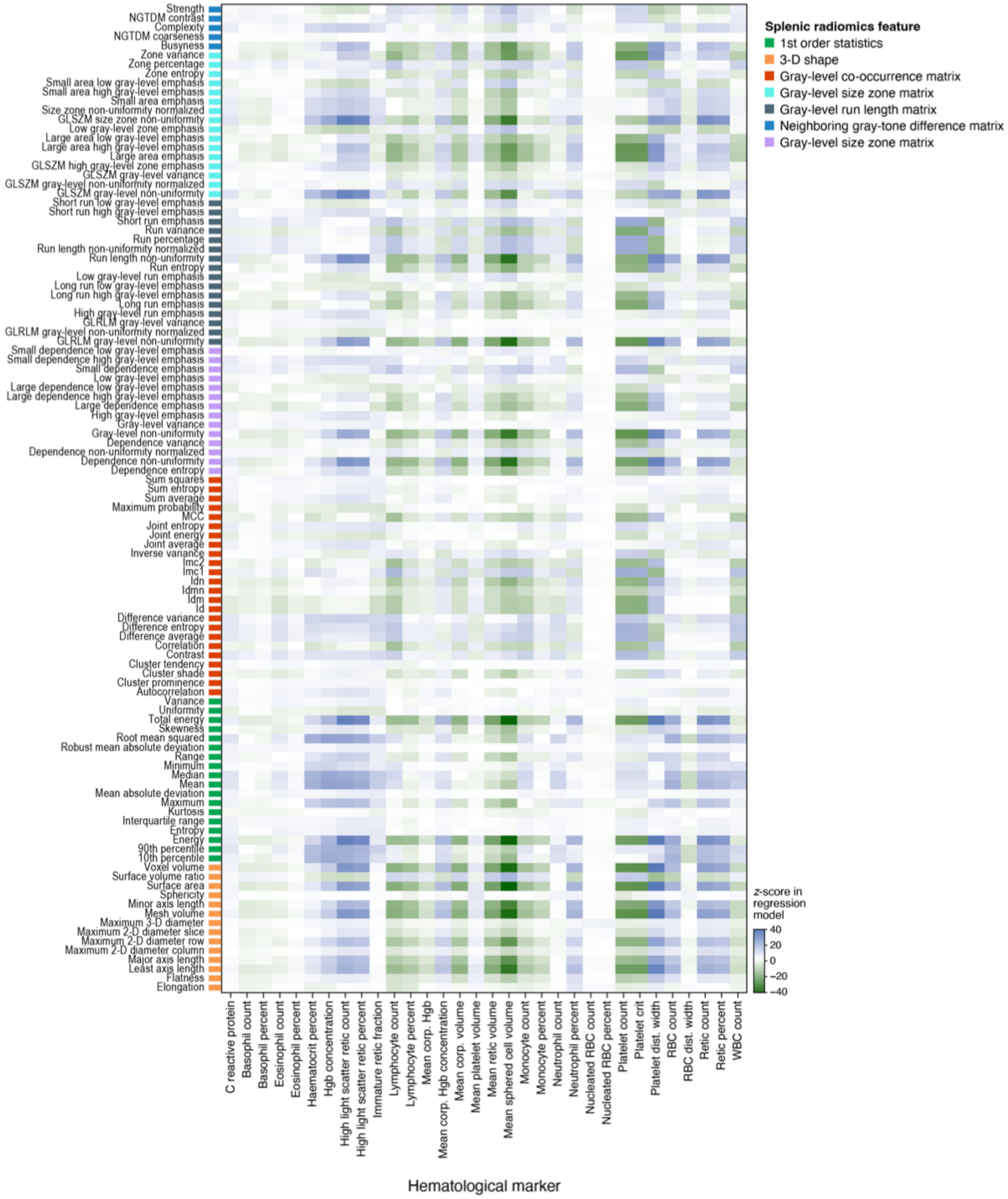

**Supplemental Figure 5.** Example Spleen Images for Radiomics Features associated with CAD. We include a standardized view of the spleens for low and high values for the ten radiomics features associated with CAD. After ordering the spleens based on lowest to highest values for each feature, examples in the low, low-middle, middle-high, and high categories correspond to an image from the lowest 500, middle 12,000 - 13,000, middle 32,000 - 33,000, and highest 500 values. Reproduced by kind permission of UK Biobank ©

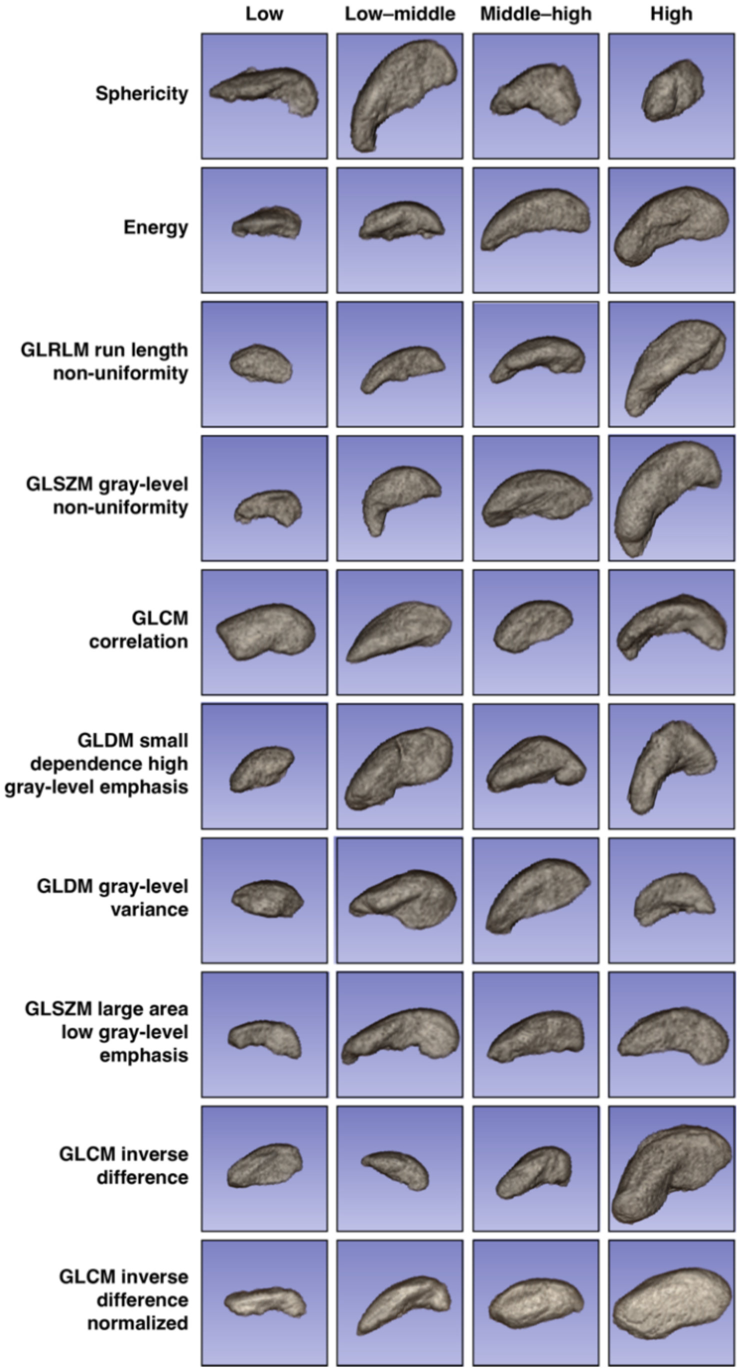

**Supplemental Figure 6.** Manhattan plot for splenic gray level size zone matrix (GLSZM) gray level non-uniformity with top 10 nearest genes annotated.

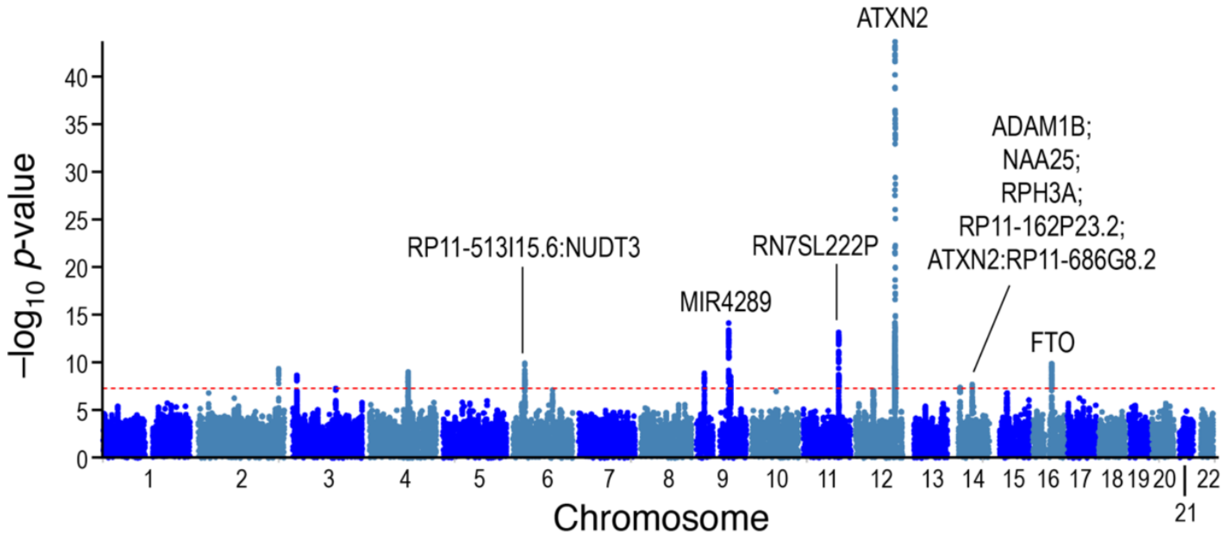

**Supplemental Figure 7.** Manhattan plot for splenic sphericity with top 10 nearest genes annotated.

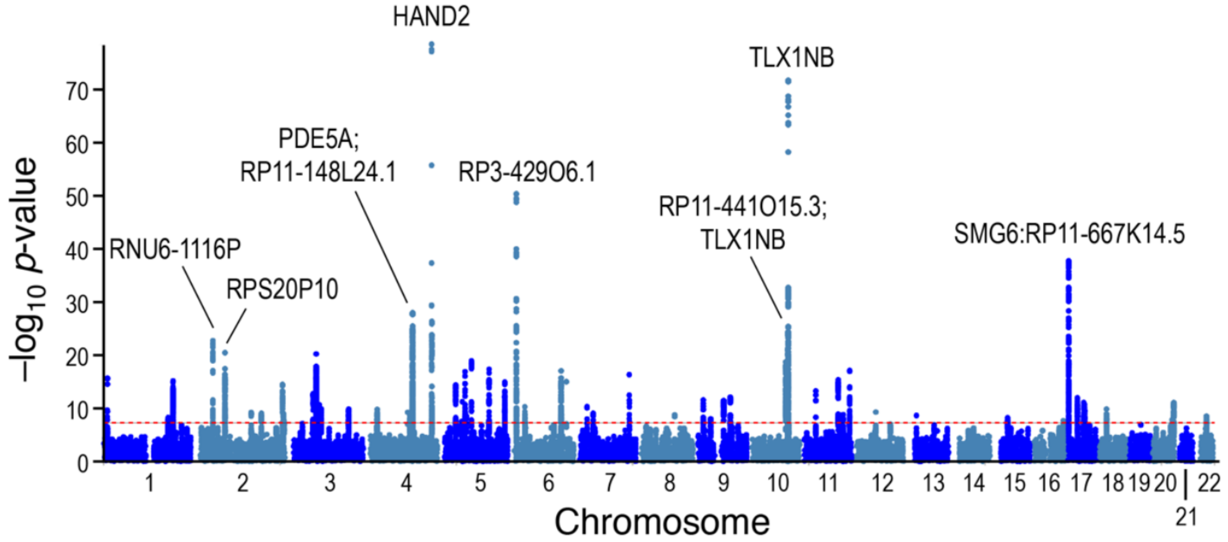

**Supplemental Figure 8.** Manhattan plot for splenic gray level co-occurrence matrix (GLCM) correlation with the nearest genes annotated.

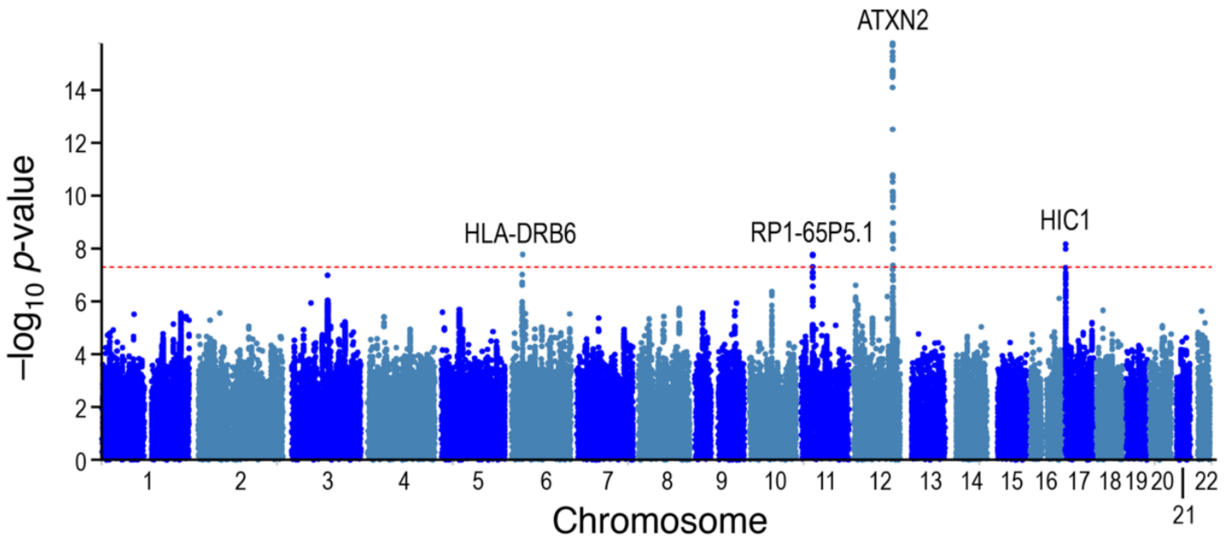

**Supplemental Figure 9.** Manhattan plot for splenic first order energy with top 10 nearest genes annotated.

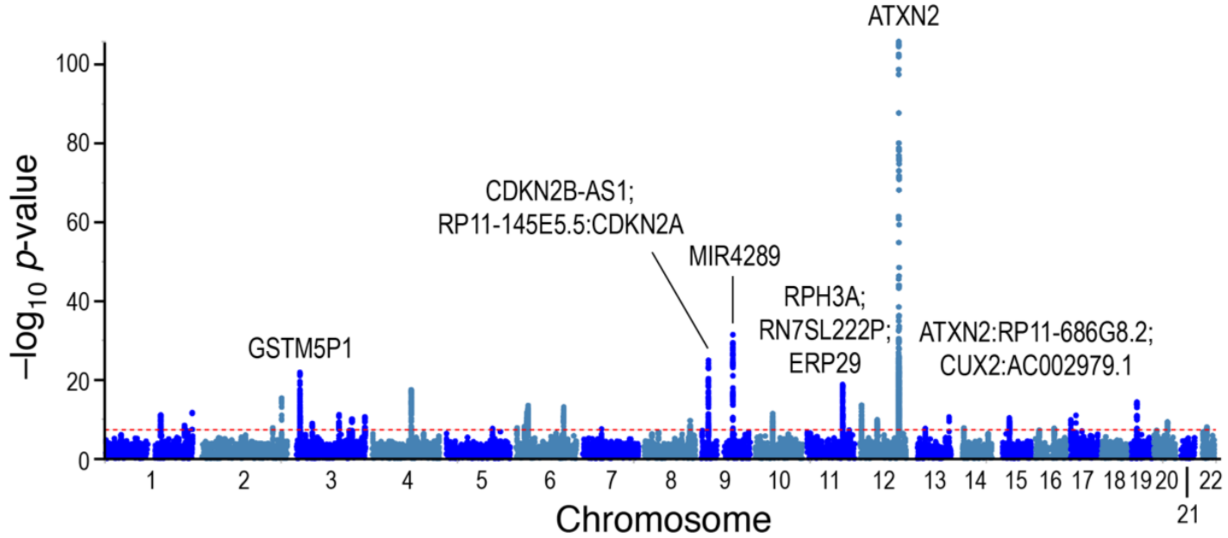

**Supplemental Figure 10.** Manhattan plot for splenic gray level dependence matrix (GLDM) small dependence high gray level emphasis with the nearest genes annotated.

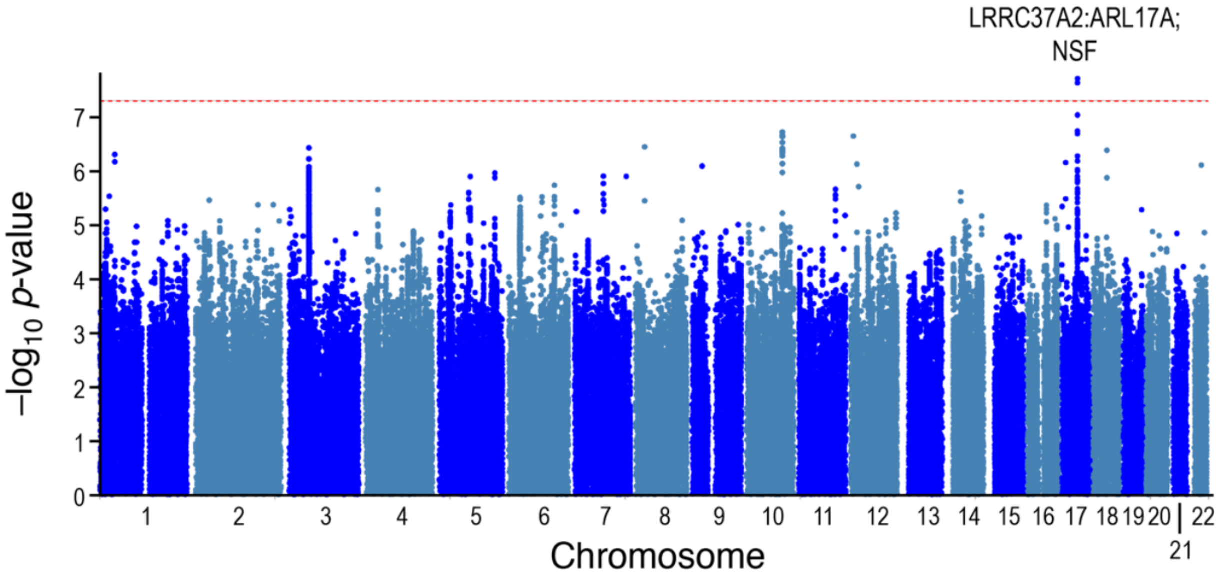

**Supplemental Figure 11.** Manhattan plot for splenic gray level size zone matrix (GLSZM) large area low gray level emphasis with top 10 nearest genes annotated.

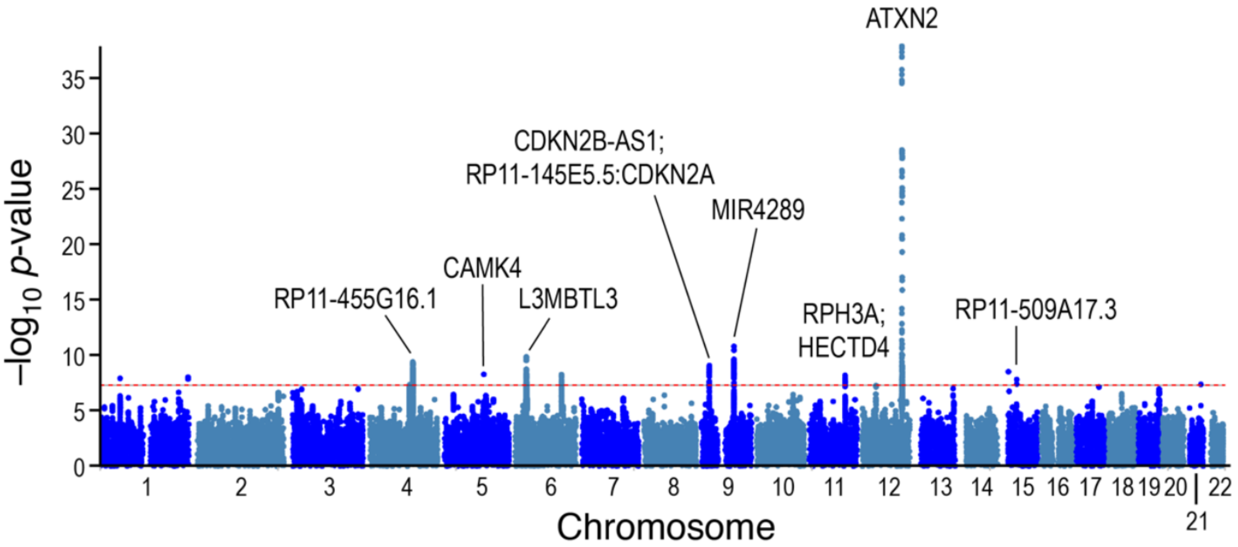

**Supplemental Figure 12.** Manhattan plot for splenic gray level run length matrix (GLRLM) run-length non-uniformity with top 10 nearest genes annotated.

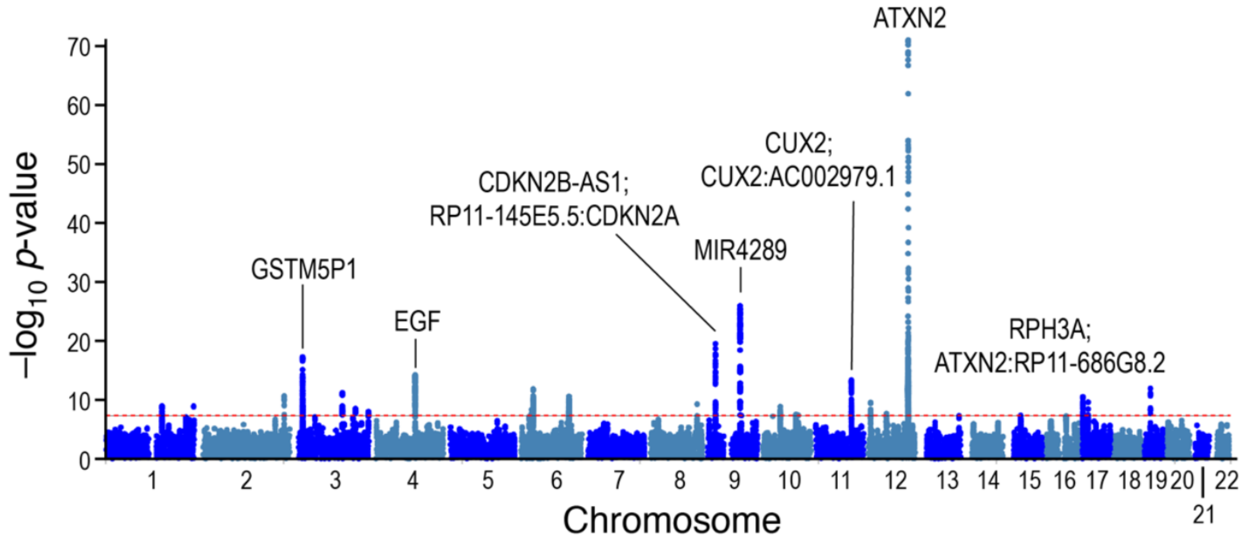

**Supplemental Figure 13.** Manhattan plot for splenic gray level co-occurrence matrix (GLCM) inverse difference with the nearest genes annotated.

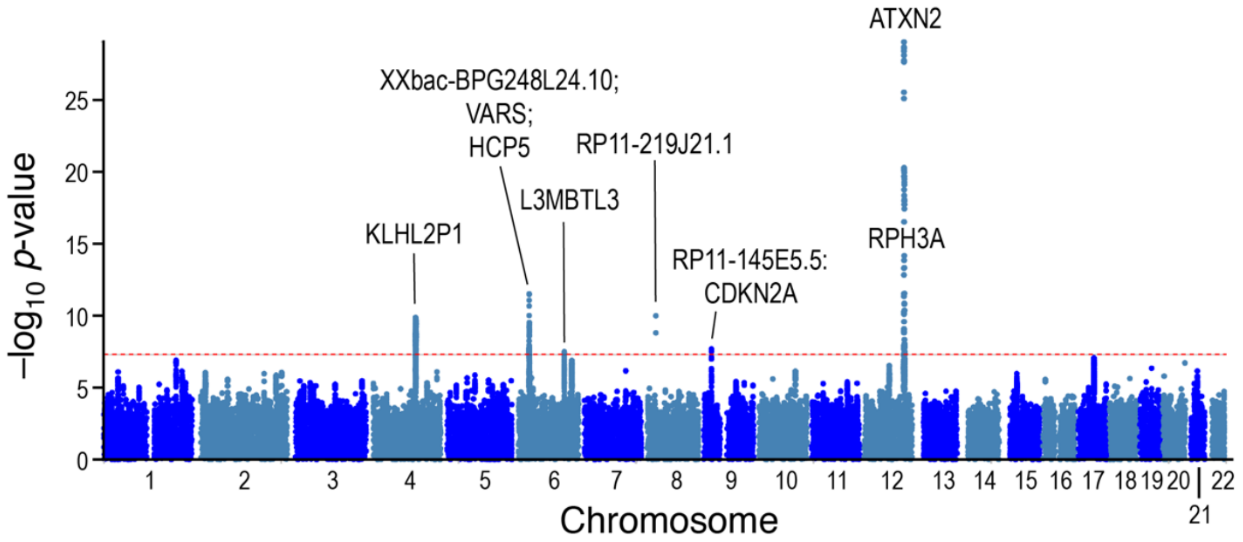

**Supplemental Figure 14.** Manhattan plot for splenic gray level co-occurrence matrix (GLCM) inverse difference normalized with the nearest genes annotated.

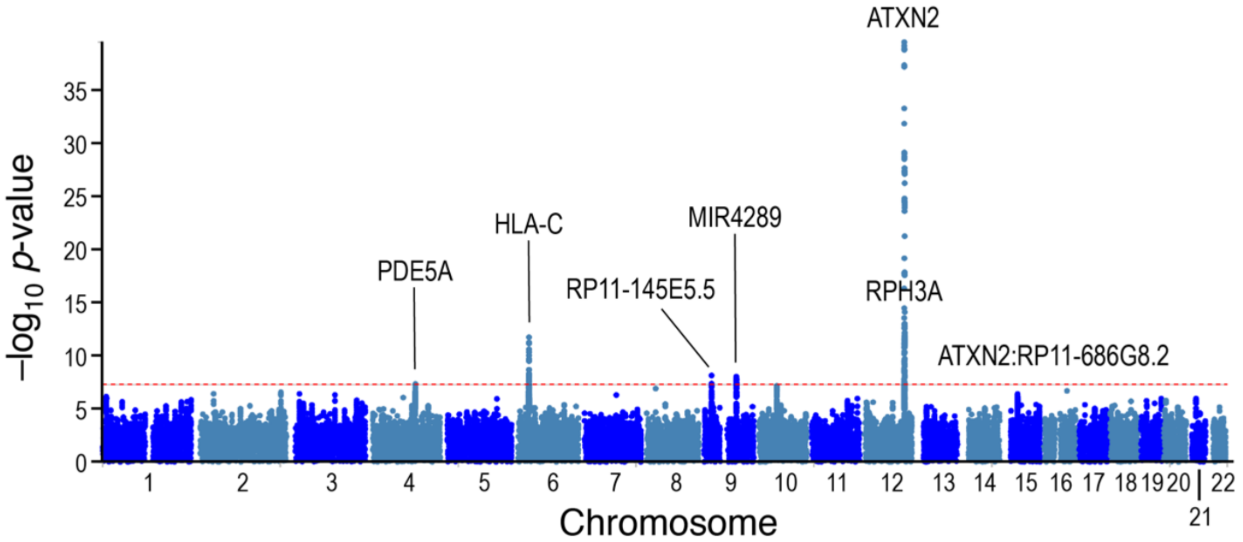

**Supplemental Figure 15.** Overlapping SNPs between CAD and splenic phenotypes and their phenotypic associations.

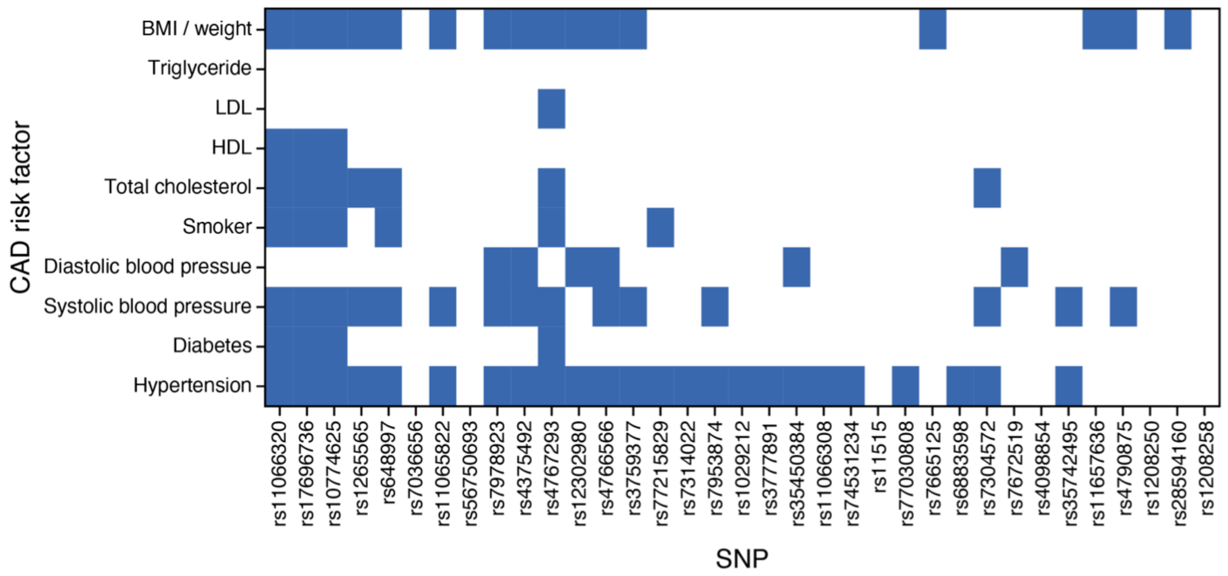
